# Clinical Implications of Restriction with Normal Spirometry: A Retrospective Cohort Study

**DOI:** 10.64898/2025.12.15.25342141

**Authors:** Alexander T. Moffett, Aparna Balasubramanian, Benjamin E. Schmid, Meredith C. McCormack, Gary E. Weissman

**Affiliations:** 1Division of Pulmonary, Allergy, and Critical Care Medicine, Department of Medicine, University of Pennsylvania, Philadelphia, PA, USA; Palliative and Advanced Illness Research (PAIR) Center, University of Pennsylvania, Philadelphia, PA, USA; Leonard Davis Institute of Health Economics, University of Pennsylvania, Philadelphia, PA, USA; Division of Pulmonary and Critical Care Medicine, Department of Medicine, Johns Hopkins University, Baltimore, MD, USA; Department of Biostatistics, Epidemiology, and Informatics, University of Pennsylvania, Philadelphia, PA, USA

**Keywords:** pulmonary function test interpretation, restriction, spirometry

## Abstract

**Background:** Though the European Respiratory Society and American Thoracic Society technical standard on pulmonary function test interpretation recommends the use of a normal forced vital capacity (FVC) to infer the absence of restriction, recent data suggest that restriction may be present in as many as one in five patients with normal spirometry. The clinical implications of restriction with normal spirometry are unknown.

**Methods:** We interpreted pulmonary function tests (PFTs) with both static and dynamic lung volume measurements performed between 2012 and 2025 at four pulmonary diagnostic labs. We used multivariable logistic regression to identify clinical characteristics associated with restriction among patients with normal spirometry and used a Cox proportional hazards model to assess the association of restriction with survival, adjusting for age, height, sex, forced expiratory volume in 1 second (FEV1) z-score, FVC z-score, and FEV1/FVC z-score.

**Results:** We interpreted 83,886 PFTs from 47,597 patients (mean age 58.8 years, 59.8% female, 63.6% White). Normal spirometry was present in 44,246 (52.7%) PFTs. Restriction was present in 11,361 (25.7%) PFTs with normal spirometry. Among patients with normal spirometry, restriction was associated with interstitial lung disease (adjusted odds ratio 3.40, 95% confidence interval [CI] 3.20–3.60) and with increased all-cause mortality (adjusted hazard ratio 1.45, 95% CI 1.34–1.57).

**Interpretation:** Restriction with normal spirometry is associated with interstitial lung disease and with increased mortality. Normal spirometry performs poorly as a screening test to exclude clinically significant ventilatory impairments.

**What Is Already Known on this Topic:** The approach to pulmonary function test interpretation recommended by the European Respiratory Society and the American Thoracic Society uses a normal forced vital capacity to infer the absence of restriction. However, recent data suggest that restriction may be more common among pulmonary function tests with normal spirometry than has been assumed.

**What this Study Adds:** This study offers the first assessment of the clinical implications of restriction in patients with normal spirometry. We found that in pulmonary function tests with normal spirometry, the presence of restriction is strongly associated with both interstitial lung disease and higher all-cause mortality.

**How this Study Might Affect Research, Practice, or Policy:** The current recommended approach to pulmonary function test interpretation fails to identify clinically significant ventilatory impairments in as many as one in four patients with normal spirometry, suggesting the need to reconsider how normal spirometry is used to inform clinical decision making.

## Introduction

A central purpose of pulmonary function test (PFT) interpretation is to determine the presence or absence of restriction.^1,2^ The European Respiratory Society and American Thoracic Society (ERS/ATS) technical standard on pulmonary function test (PFT) interpretation defines restriction by a total lung capacity (TLC) less than the lower limit of normal (LLN).^3^ According to this technical standard, while the measurement of static lung volumes is necessary to determine the presence of restriction, the absence of restriction can be inferred from spirometry alone if the forced vital capacity (FVC) is greater than the LLN.^3,4^

In support of this recommendation, the ERS/ATS technical standard cites a single study in which the negative predictive value (NPV) of the FVC LLN was estimated at 97.6%.^5^ This study, however, was performed at a single site, was limited to White patients, involved fewer than two thousand PFTs, and employed race-specific reference equations that are no longer recommended by ERS/ATS.^6^ While other single-site studies using race-specific reference equations have reported similar NPV estimates,^7–13^ a recent multi-site study using modern race-neutral reference equations found the NPV to be substantially lower, at only 80.5%.^14^ This more recent estimate suggests that interpreting PFTs in accordance with the ERS/ATS technical standard may falsely exclude restriction in almost 1 in 5 PFTs with a normal FVC.

Though the prevalence of restriction with normal spirometry appears greater than previously assumed, the clinical implications of this pattern are unknown. If restriction with normal spirometry is associated with an increased risk of respiratory disease and adverse outcomes, the ERS/ATS approach to PFT interpretation may contribute to delays in diagnosis and treatment Conversely, if no such association exists, restriction with normal spirometry may instead represent an artifact resulting either from differences in the reference equations used to define the FVC and TLC LLNs,^15,16^ or from variation resulting from the distinct technical maneuvers employed in FVC and TLC measurement.^17–19^ To address this gap, we performed a retrospective cohort study to describe the clinical characteristics and events associated with restriction in PFTs with normal spirometry.

## Study Design and Methods

### Study Design

This retrospective cohort study was performed in accordance with the Strengthening the Reporting of Observational Studies in Epidemiology (STROBE) guidelines for reporting observational studies (**e-Table 1**).^20^ The University of Pennsylvania Institutional Review Board granted an exemption for this study.

### Study Population

We included PFTs with both static and dynamic lung volume measurements that were performed between January 2012 and July 2025 at one of four pulmonary diagnostic labs within a single university health system. Patient age, height, sex, body mass index (BMI), and pack-years were as documented on the PFT report. Patient race and ethnicity were as documented in the electronic health record (EHR). We excluded PFTs with missing demographic or anthropometric data, as well as those from patients less than 18 years of age or more than 80 years of age at the time of testing. We followed patients from their first encounter in the electronic health record until July 2025.

### Pulmonary Function Tests

The PFTs included in this study were performed as part of routine clinical practice in accordance with ATS recommendations. Static lung volumes were measured with plethysmography. PFTs were interpreted in accordance with the current ERS/ATS technical standard.^3^ The forced expiratory volume in 1 second (FEV1), FVC, and FEV1/ FVC z-scores were calculated with Global Lung Function Initiative (GLI) Global reference equations.^16^ TLC z-scores and DLCO z-scores were calculated with GLI equations.^15,21^

A PFT parameter was defined as abnormal if its z-score was less than −1.645, and as normal otherwise. Restriction was defined by an abnormal TLC. Restriction with normal spirometry was defined by the presence of restriction along with a normal FVC and a normal FEV1/FVC.

### Clinical Characteristics

For each PFT we determined the presence or absence of each of the following clinical characteristics: cough, dyspnea, wheezing, asthma, bronchiectasis, chest wall disorder, chronic obstructive pulmonary disease (COPD), interstitial lung disease (ILD), neuromuscular disorder, bronchial wall thickening, emphysema, honeycombing, reticulation, traction bronchiectasis, and technical adequacy of spirometry performance.

Respiratory symptoms were assessed at the time of pulmonary function testing and documented in structured fields in the PFT report. Cough was present if the patient endorsed a productive or non-productive cough. Dyspnea was present if the patient endorsed dyspnea on hills and stairs, with walking, or with any exertion. Wheezing was present if the patient endorsed rare, frequent, or constant wheezing.

Respiratory diseases were assessed by applying multimodal automated phenotyping (MAP)—an unsupervised method that has been previously validated for asthma and COPD in a similar clinical population—to the complete EHR data for each patient.^22^ Respiratory disease types were drawn from the 2005 ATS/ERS guidelines for PFT interpretation.^23^ For each patient and each respiratory disease, we calculated the number of outpatient clinical encounters for the patient in which an International Classification of Disease (ICD) code associated with the disease was assigned (**e-Table 2**), the total number of references to the disease in the patient’s outpatient progress notes (**e-Table 3**), and the total number of outpatient encounters for the patient. With these inputs, the MAP algorithm was then used to predict the presence or absence of each respiratory disease for each patient.^22^

CT findings were assessed by applying natural language processing (NLP) with regular expressions to determine whether, for each patient, a chest computed tomograph (CT) was performed in which bronchial wall thickening, emphysema, honeycombing, reticulation, or traction bronchiectasis were referenced in the impression or narrative of the radiology report (**e-Table 4**).^24^

The technical adequacy of spirometry data was assessed by applying NLP to the comments of respiratory technicians to classify patient effort as adequate or inadequate (**e-Table 5**).

Logistic regression was used to compare the odds of the above clinical characteristics among PFTs with normal spirometry with and without restriction. Analyses were adjusted for age, height, sex, FEV1 z-score, FVC z-score, and FEV1/FVC z-score. Subgroup analyses were performed by sex, race, pulmonary diagnostic lab, and the medical specialty of the referring physician.

In a secondary analysis, we compared the odds of these clinical characteristics in PFTs with restriction with and without normal spirometry, adjusting for patient age, sex, height, and TLC z-score.

### Events

Cox proportional hazards models were used to compare the time from PFT performance to an emergency department (ED) visit with a respiratory complaint (**e-Table 6**) and to mortality from any cause, in PFTs with normal spirometry, with and without restriction. Analyses were adjusted for age, sex, height, FEV1 z-score, FVC z-score, and FEV1/FVC z-score.

### Repeat Testing

To assess the persistence of restriction with normal spirometry on repeat testing, we identified patients with this interpretation on their initial PFT, and compared this interpretation to that of all of their subsequent PFTs. We also identified all PFTs with follow up testing and calculated the prevalence of pairs of interpretations on initial and repeat testing.

### Sensitivity Analysis

To assess the sensitivity of our findings to the use of the MAP algorithm, we repeated our analysis defining a respiratory disease as present if the EHR data for a patient included at least one ICD-10 code associated with the disease. To assess the sensitivity of our findings to the adoption of race-neutral reference equations, we repeated our analysis with the race-specific GLI 2012 reference equations.^25^

### Statistical Computing

The analysis was performed with R version 4.5.1. The MAP algorithm was implemented with the MAP package.^26^ All statistical tests were two sided and a *P* value < 0.05 was interpreted as statistically significant. All code is open source and publicly accessible (https://github.com/weissman-lab/restriction-with-normal-spirometry).

## Results

We interpreted 83,886 PFTs from 47,597 patients (**Table 1**, **e-Figure 1**). Most PFTs were from female (50,196 [59.8%]) and from White (50,859 [63.6%]) or Black (23,495 [29.4%]) patients. The mean age was 58.8 years (standard deviation [SD] 14.0). The mean FVC, FEV1/FVC, and TLC z-scores were −1.0 (SD 1.4), −0.7 (SD 1.4), and −1.3 (SD 1.6), respectively. FVC and TLC z-scores were lower among male patients, non-White patients, patients tested at pulmonary diagnostic labs 1 and 2, and patients referred for pulmonary function testing by a pulmonologist (**e-Tables 7–10**). The median follow-up time was 32 (interquartile range 63) months. Pack years, symptoms, imaging, and bronchodilator response data were missing for a significant fraction of PFTs (**e-Table 11**).

**Table 1:**
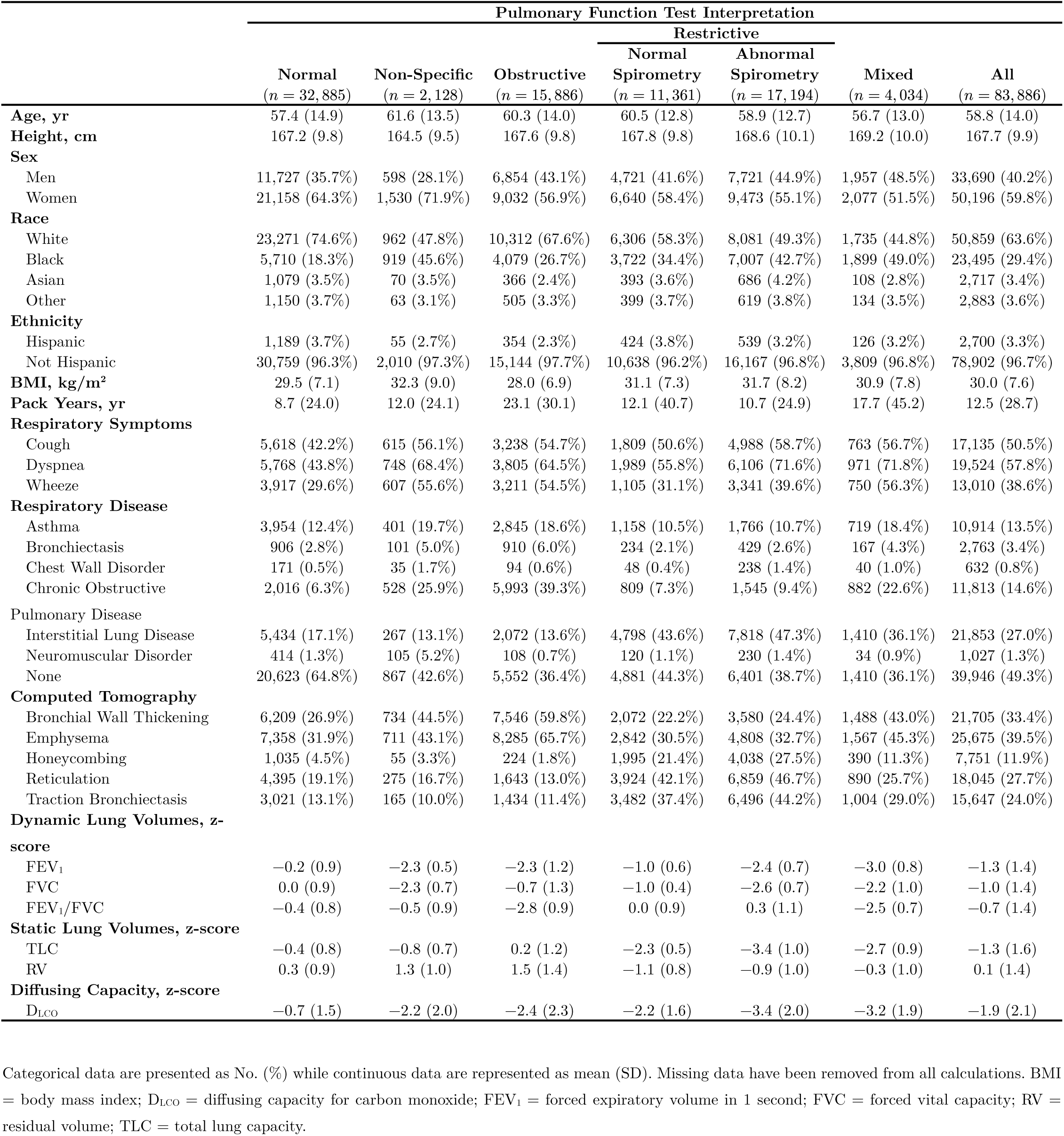
Clinical and Demographic Characteristics.

Normal spirometry was present in 44,246 (52.7%) PFTs. Among PFTs with normal spirometry, the prevalence of restriction was 25.7% (95% CI 25.3–26.1) (**Figure 1**). Restriction of any form was present in 32,589 (38.8%) PFTs. Restriction with normal spirometry was present in 11,361 (13.5%) PFTs, while restriction with abnormal spirometry was present in 17,194 (20.5%) PFTs, and a mixed ventilatory impairment was present in 4,034 (4.8%) PFTs. The median TLC z-score among PFTs with restriction with normal spirometry was −2.2 (first quartile −2.6, third quartile −1.9) (**Figure 2**).

**Figure 1.**
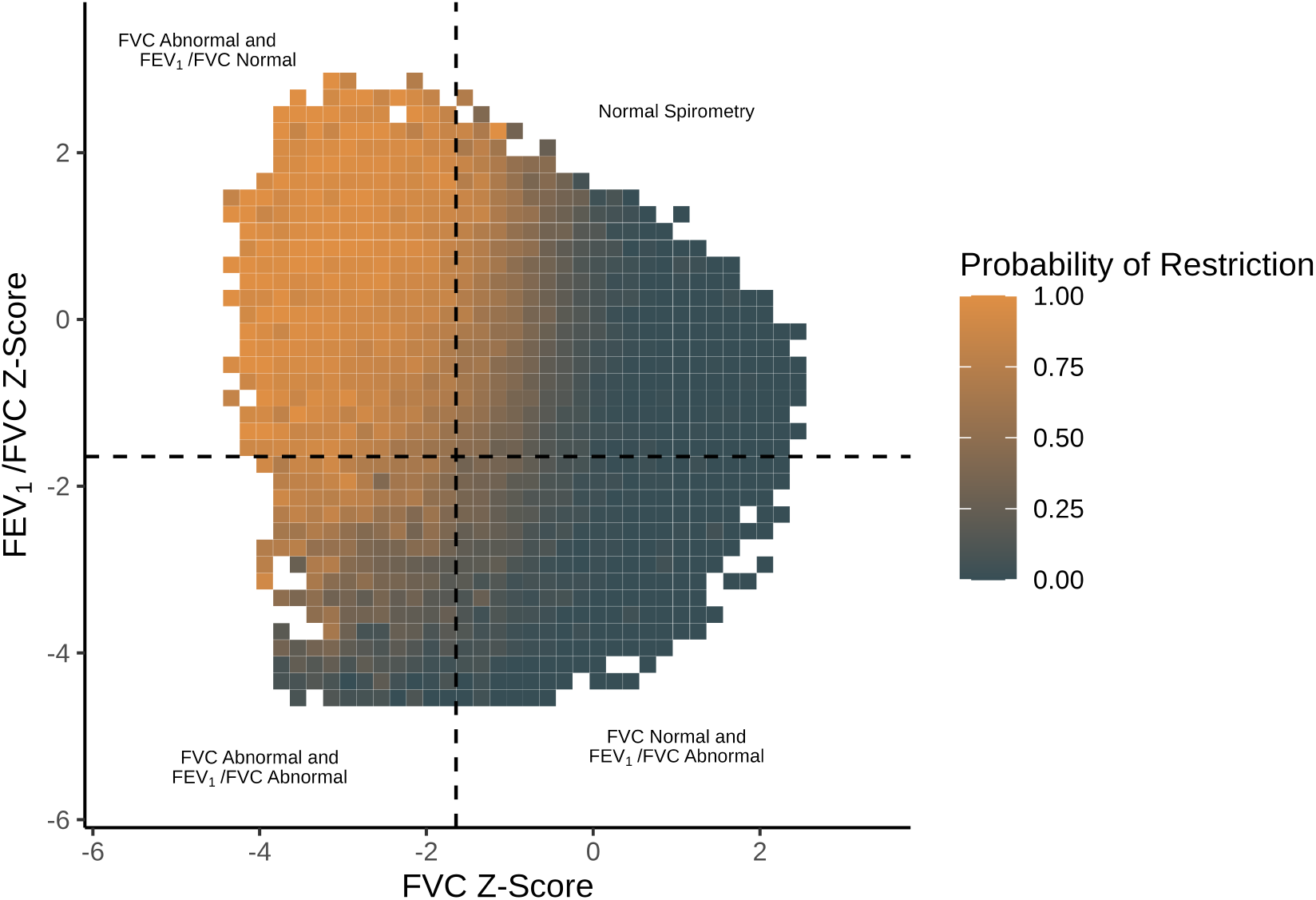
The association between FVC and FEV1/FVC z-scores and restriction. Data are drawn from 83,886 PFTs performed at four pulmonary diagnostic labs between 2012 and 2025. Restriction is present if the TLC z-score is less than −1.645. Data are binned by z-score intervals of 0.2. The probability of restriction for a given bin is equal to the proportion of PFTs within the bin in which restriction is present. FVC and FEV1/FVC z-scores are calculated using GLI Global reference equations while TLC z-scores are calculated using GLI 2019 equations. FEV1 = forced expiratory volume in 1 second; FVC = forced vital capacity; GLI = Global Lung Function Initiative; PFT = pulmonary function test; TLC = total lung capacity.

**Figure 2.**
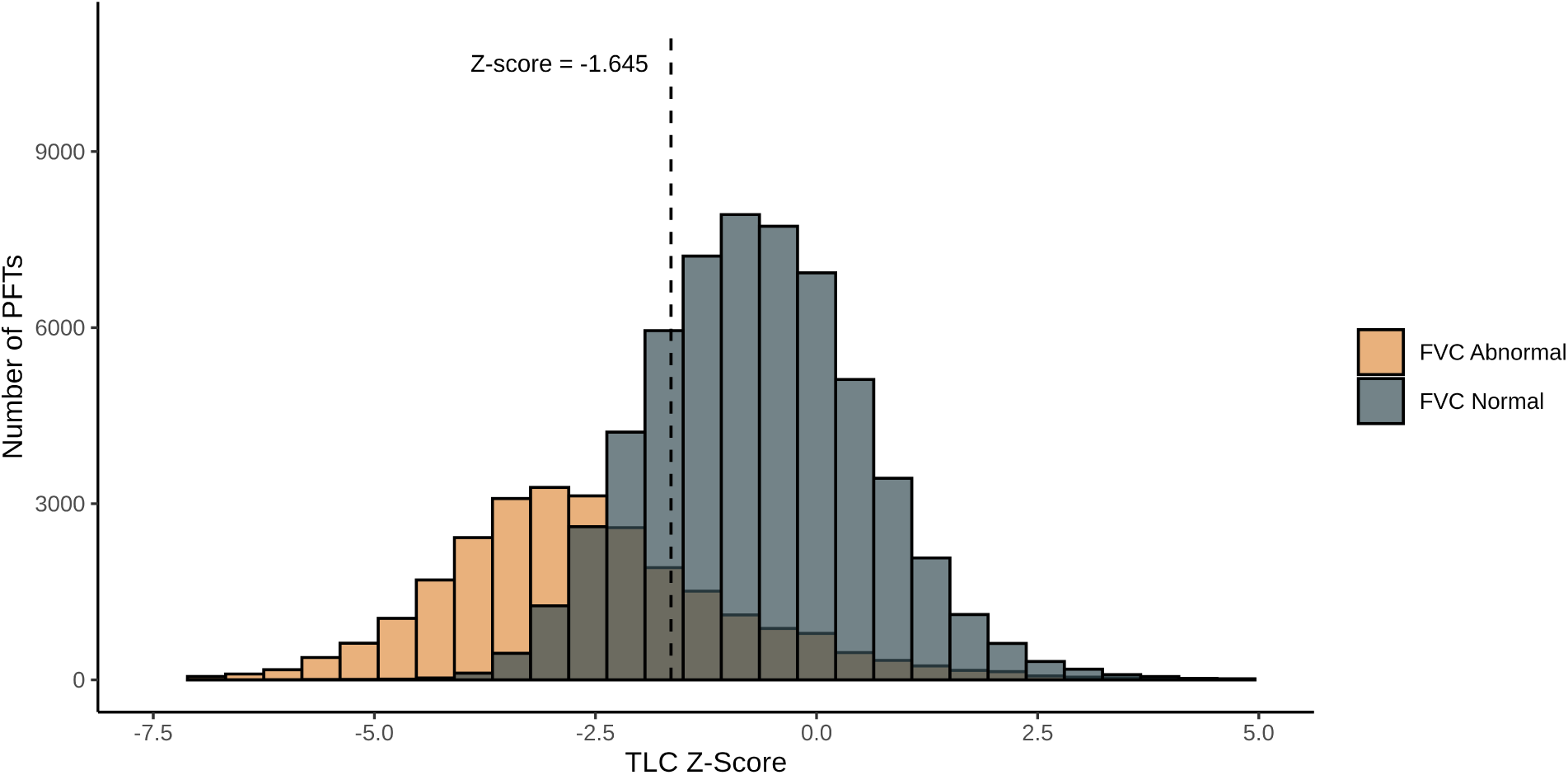
Distribution of TLC z-scores among patients with normal and abnormal FVCs. Data are drawn from 83,886 PFTs performed at four pulmonary diagnostic labs between 2012 and 2025. FVC = forced vital capacity; PFT = pulmonary function test; TLC = total lung capacity.

Among PFTs with normal spirometry, restriction was more likely in older patients (adjusted odds ratio [aOR] 1.01 per year, 95% CI 1.01–1.01), in more obese patients (aOR 1.00 per kg/m^2^, 95% CI 1.00–1.01), in Black patients (aOR 1.34, 95% CI 1.26–1.43), and in patients with longer smoking histories (aOR 1.00 per pack-year, 95% CI 1.00–1.00) (**e-Table 12**). Among PFTs with normal spirometry, restriction was less likely in female patients (aOR 0.56, 95% CI 0.52–0.61), in non-Hispanic patients (aOR 0.83, 95% CI 0.72–0.96), in patients with a positive bronchodilator response (aOR 0.39, 95% CI 0.28–0.52) and in patients with a greater residual volume (RV) (aOR 0.01, 95% CI 0.01–0.01) or a greater DLCO (aOR 0.61, 95% CI 0.60–0.62). Restriction was not associated with adequacy of effort on spirometry (aOR 0.83, 95% CI 0.46–1.56)

Among PFTs with normal spirometry, restriction was associated with an increased probability of chronic cough (aOR 1.25, 95% CI 1.15–1.36) and dyspnea (aOR 1.20, 95% CI 1.10–1.31), with the diagnosis of ILD (aOR 3.40, 95% CI 3.20–3.60) and with honeycombing (aOR 4.47, 95% CI 4.03–4.96), reticulation (aOR 2.63, 95% CI 2.26–2.81), and traction bronchiectasis (aOR 3.48, 95% CI 3.24–3.74) (**Figure 3**). Restriction was associated with a reduced probability of wheezing (aOR 0.87, 95% CI 0.79–0.96), asthma (aOR 0.79, 95% CI 0.73–0.86), bronchiectasis (aOR 0.59, 95% CI 0.50–0.69), a chest wall disorder (aOR 0.68, 95% CI 0.47–0.97), COPD (aOR 0.82, 95% CI 0.74–0.90), a neuromuscular disorder (aOR 0.68, 95% CI 0.54–0.86), bronchial wall thickening (aOR 0.67, 95% CI 0.63–0.72), and emphysema (aOR 0.79, 95% CI 0.74–0.84). Restriction was associated with an increased risk of an emergency department visit with a respiratory complaint (adjusted hazard ratio [aHR] 1.18, 95% CI 1.11–-1.26) and with increased all-cause mortality (aHR 1.45, 95% CI 1.34–-1.57) (**Figure 4**). Results were generally similar among patients stratified by sex, race, pulmonary diagnostic laboratory, and referring provider specialty (**e-Tables 13–16**), though statistically significant associations with mortality were not seen in patients of Asian or other races, or in patients tested at pulmonary diagnostic labs 2 and 4.

**Figure 3.**
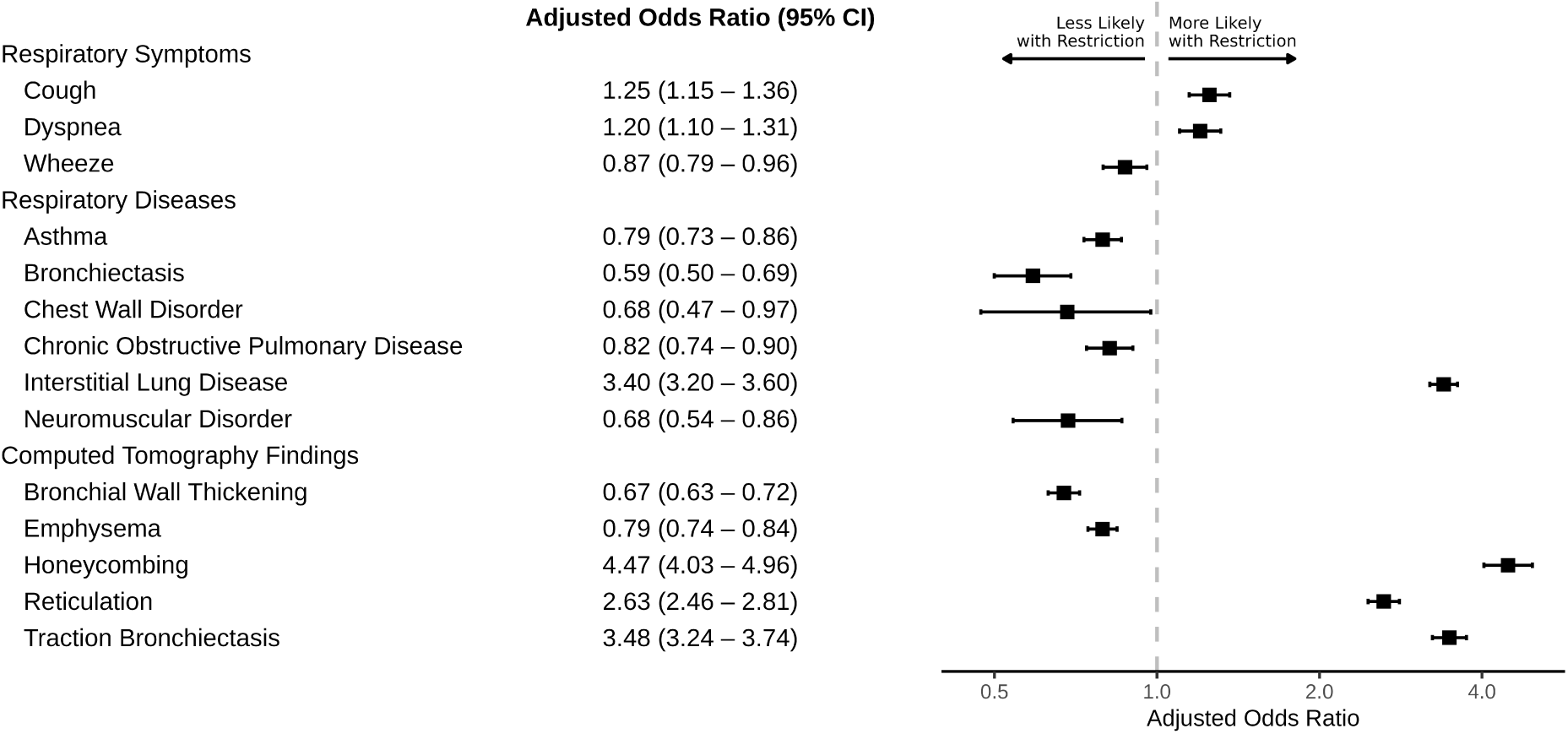
Outcomes associated with restriction in pulmonary function tests with normal spirometry. Logistic regression models are used to predict respiratory symptoms, respiratory diseases, and chest computed tomography findings on the basis of restriction, adjusting for age, sex, height, FEV1 z-score, FVC z-score, and FEV1/FVC z-score. Data are drawn from 44,246 PFTs with normal spirometry performed at four pulmonary diagnostic labs between 2012 and 2025.

**Figure 4.**
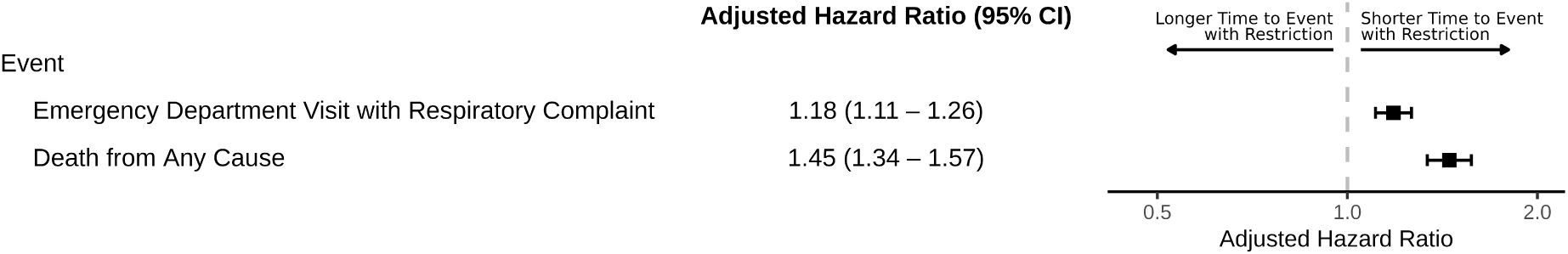
Events associated with restriction in PFTs with normal spirometry. Cox proportional hazards models are used to predict the time from PFT performance to an emergency department visit with respiratory complaint and time to death from any cause on the basis of restriction, adjusting for age, sex, height, FEV1 z-score, FVC z-score, and FEV1/ FVC z-score. Data are drawn from 44,246 PFTs with normal spirometry performed at four pulmonary diagnostic labs between 2012 and 2025.

Among PFTs with restriction, patients with normal spirometry were more likely than those with abnormal spirometry to have ILD (aOR 1.95, 95% CI 1.84–2.07) or to have evidence of honeycombing (aOR 1.54, 95% CI 1.43–1.66), reticulation (aOR 1.79, 95% CI 1.68–1.90), or traction bronchiectasis (aOR 1.56, 95% CI 1.46–1.66) (**e-Table 17**).

Restriction with normal spirometry was present on the initial PFTs of 5,346 patients and was still present on 922 (54.0%, 95% CI 51.6–56.4), 539 (55.3%, 95% CI 52.1–58.4), 379 (54.1%, 95% CI 50.4–57.9), and 273 (54.4%, 95% CI 49.9–58.8) patients on their second, third, fourth, and fifth PFTs, respectively (**Figure 5a**). Among PFTs with follow up testing, restriction with normal spirometry was present in 3,670 (62.9%, 95% CI 61.7–64.2) tests, while 1,095 (18.8%, 95% CI 17.8–19.8) were normal, 792 (13.6%, 95% CI 12.7–14.5) were restrictive with abnormal spirometry, and 153 (2.6%, 95% CI 2.2–3.1) tests were mixed (**Figure 5b**).

**Figure 5.**
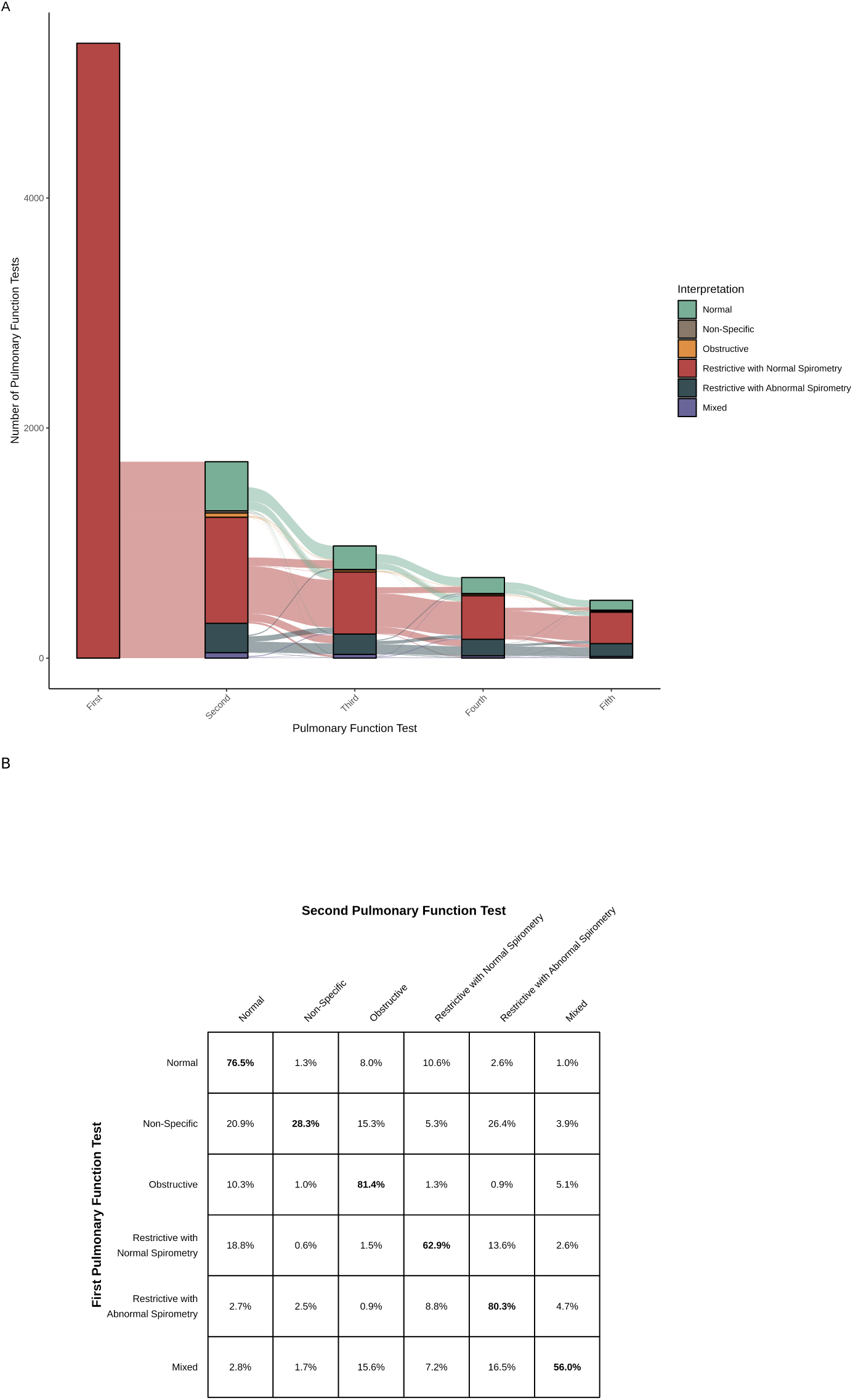
Changes in PFT interpretation on repeating testing. (A) An alluvial plot displays the PFT interpretations for 5,346 patients with restriction with normal spirometry present on their initial PFT. Strata within each axis represent the PFT interpretations for these patients on repeat testing. Alluvia between axes represent the interpretation of the preceding PFT. (B) A grid displays the probability of the PFT interpretation of a second PFT as a function of the PFT interpretation of the first PFT for 36,289 repeated PFTs. Rows display the interpretation of the first PFT while columns display the interpretation of the second PFT. PFT = pulmonary function test.

In our sensitivity analyses, defining respiratory disease diagnoses by the presence of at least one associated ICD-10 code did not significantly affect our findings (**e-Table 18**) Race-specific reference equations did not have a significant impact on the prevalence of restriction with normal spirometry (25.7% versus 25.5%), on its persistence on repeat testing (**e-Figure 2**), or on its association with respiratory symptoms, diseases, CT findings, or events (**e-Tables 19–20**).

## Discussion

In this retrospective cohort study involving more than 80,000 PFTs performed at four pulmonary diagnostic labs, restriction was present in more than 1 in 4 PFTs with normal spirometry, and was associated with both ILD and increased mortality. These findings suggest that restriction with normal spirometry is clinically significant, and that normal spirometry performs poorly as a screening test for clinically significant ventilatory impairments.

Our study challenges the approach to PFT interpretation recommended by the ERS/ ATS technical standard, in which the presence of a normal FVC is used to infer the absence of restriction.^3,4^ In our cohort, the prevalence of restriction among patients with normal spirometry was 25.7%, more than an order of magnitude greater than the prevalence of 2.4% cited by ERS/ATS in support of the current recommended interpretive approach.^5^ While many of the PFTs included in this study were also included in our prior multisite study,^14^ the prevalence of restriction with normal spirometry among those not included was 31.7%, suggesting that our prior study may have underestimated the prevalence of this finding. In our cohort in the present study, the use of a normal FVC to exclude restriction would have resulted in the misclassification of more than 1 in 4 PFTs with normal spirometry and the failure to identify more than 1 in 3 PFTs with restriction.

Moreover, our study suggests that such misclassification has important clinical implications, as restriction with normal spirometry is associated with distinct clinical characteristics and events. Among patients with normal spirometry, those with restriction were more likely to be diagnosed with ILD and to have evidence of honeycombing, reticulation, or traction bronchiectasis on chest CT. They were less likely to be diagnosed with asthma or COPD and less likely to have evidence of bronchial wall thickening or emphysema on chest CT. And they had higher rates of ED encounters with respiratory complaints and of all-cause mortality. By classifying patients both with and without restriction as having normal spirometry, the approach to PFT interpretation recommended by ERS/ATS obscures important clinical heterogeneity which may contribute to delays in the diagnosis and treatment of respiratory disease.

Perhaps surprisingly, in our secondary analysis we found that among patients with restriction, those with normal spirometry were more likely to have ILD than those with abnormal spirometry. It is not simply that respiratory disease is possible in patients with normal spirometry,^27–31^ but instead that normal spirometry can itself be a sign of such disease.

Three mechanisms may contribute to restriction with normal spirometry. First, restriction with normal spirometry may result from a reduced RV, as is seen, for instance, in fibrotic ILD.^32^ As TLC is equal to the sum of FVC and RV, a reduction in RV can produce an abnormal TLC despite a normal FVC.^33^ In our cohort, the RV was significantly lower in patients with restriction with normal spirometry than in patients with normal spirometry without restriction, with a z-score of −1.1 in the former and 0.3 in the latter. In this respect, restriction with normal spirometry can be seen as the obverse of the non-specific pattern, in which the FVC is abnormal, while the FEV1/FVC and TLC are normal, allowing for the combination of an abnormal FVC and a normal TLC.^34^

Second, restriction with normal spirometry may stem from differences in the cohorts used to define FVC and TLC reference equations.^6,35^ While the GLI Global equations for spirometry are based on a global cohort of healthy White, Black, North and South East Asian individuals, the equations for static lung volumes are based on a cohort of healthy White individuals drawn primarily from Europe.^15,16^ If the lung function of the latter cohort were greater than that of the former—as would be expected given the effect that socioeconomic factors have on lung function even among healthy volunteers—this would lead to a discordance in the TLC and FVC LLN, resulting in a greater number of patients with both a normal FVC and an abnormal TLC than if these equations had been defined with the same cohort.^36–38^ In the absence of global race-neutral reference equations for static lung volumes, the precise contribution of such discordance to the prevalence of restriction with normal spirometry is unclear. Nonetheless, the close association between restriction with normal spirometry and ILD suggests that restriction with normal spirometry represents more than just a statistical artifact related to the use of discordant reference equations.

Third, restriction with normal spirometry may result from errors in static or dynamic lung volume measurement, as these values are obtained from different maneuvers with different assessments of technical adequacy.^17–19^ Nonetheless, if this were the primary mechanism responsible for the findings in this study, we would expect to see a greater degree of variability of interpretation on repeat pulmonary function testing. Moreover, among patients with normal spirometry, we did not find an association between restriction and the documentation of concerns regarding spirometric effort.

While our study challenges the approach to PFT interpretation recommended by ERS/ ATS, the proper response to this challenge remains to be determined. One possible response would be to incorporate restriction with normal spirometry into PFT interpretation as a distinct pattern, something analogous to preserved ratio impaired spirometry (PRISm).^39^ Another possible response would be to see the prevalence of restriction with normal spirometry as a reason to reform the current approach to PFT interpretation, for instance by increasing the FVC z-score cutoff used to define normal spirometry. The use of a fifth percentile cutoff is a convention and a more conservative approach to the identification of normal spirometry may be indicated if the current cutoff obscures the presence of clinically significant respiratory impairments. The proper response will depend upon the determination of the costs associated with failing to identify restriction and the benefits of avoiding the need for static lung volume measurement.

### Strengths and Limitations

This study has several strengths. First, we offer the first assessment of the clinical implications of restriction in patients with normal spirometry, providing initial evidence in support of its clinical significance. Second, we include the interpretation of more than 80,000 PFTs performed at four different pulmonary diagnostic labs, with subgroup analyses comparing results across sex, race, pulmonary diagnostic lab, and referring specialty. Third, we use EHR data—including both structured and unstructured data—to assess the diagnoses, radiographic findings, and events associated with restriction with normal spirometry.

This study also has important limitations. First, as our cohort was limited to PFTS with both static and dynamic lung volume measurements, there is the potential for ascertainment bias. Because patients referred for both static and dynamic lung volume measures likely have a higher baseline probability of having respiratory disease, the prevalence of restriction in this population may be higher than in the general population. However, the selection effects that would bias this study are similar to all previous studies of the association between spirometry and restriction in that they are not prospective evaluations of healthy controls. Restriction with normal spirometry is primarily a concern when pulmonary function testing includes only dynamic lung volume measurement, and differences in this patient population may limit the external validity of our findings. Second, as this was a retrospective cohort study using EHR data, our cohort had a significant amount of missing data, suggesting the potential for selection bias.^40^ Third, while we included PFTs from four pulmonary diagnostic labs, all labs were within a single health system. Moreover, prevalence of both restriction and ILD were higher in our cohort than has been reported in other studies of PFT interpretation. It will be important to confirm our findings in future prospective studies involving different cohorts.

### Interpretation

Restriction with normal spirometry is more common than has been previously reported and is associated with ILD and with decreased survival. Clinically significant ventilatory impairments are common among patients with normal spirometry.

## Data Availability

All data produced in the present study are available upon reasonable request to the authors

https://github.com/weissman-lab/restriction-with-normal-spirometry

## Contributions

ATM participated in study design and analysis, drafted and revised the manuscript, and is the guarantor of this work. AB, MCM, and GEW contributed to study design and revised the manuscript. BES assisted with data procurement and revised the manuscript. All authors have read and approved the manuscript.

## IRB Approval

The University of Pennsylvania Institutional Review Board provided an exemption for this study.

## Abbreviations

BMI: body mass index
COPD: chronic obstructive pulmonary disease
DLCO: diffusing capacity of carbon monoxide
FEV1: forced expiratory volume in 1 second
FVC: forced vital capacity
GLI: Global Lung Function Initiative
ILD: interstitial lung disease
PFT: pulmonary function test
RV: residual volume
TLC: total lung capacity.

