## Supplement for "Clinical Implications of Restriction with Normal Spirometry: A Retrospective Cohort Study"

**Tables**


| | |
|---|---|
| **e-Table 1** | STROBE Checklist |
| **e-Table 2** | Expressions in PFT Reports Associated with Adequate and Inadequate Effort |
| **e-Table 3** | ICD-10 Codes for Respiratory Diseases |
| **e-Table 4** | Expressions in Clinic Notes Associated with Respiratory Diseases |
| **e-Table 5** | Expressions in Radiographic Reports Associated with CT Findings |
| **e-Table 6** | Respiratory Reasons for ED Visits |
| **e-Table 7** | Characteristics by Patient Sex |
| **e-Table 8** | Characteristics by Patient Race |
| **e-Table 9** | Characteristics by Patient Pulmonary Diagnostic Lab |
| **e-Table 10** | Characteristics by Referring Specialty |
| **e-Table 11** | Missing Data |
| **e-Table 12** | Characteristics Associated with Restriction in PFTs with Normal Spirometry |
| **e-Table 13** | Outcomes Associated with Restriction in PFTs with Normal Spirometry, by Sex |
| **e-Table 14** | Outcomes Associated with Restriction in PFTs with Normal Spirometry, by Race |
| **e-Table 15** | Outcomes Associated with Restriction in PFTs with Normal Spirometry, by Pulmonary Diagnostic Lab |
| **e-Table 16** | Outcomes Associated with Restriction in PFTs with Normal Spirometry, by Referring Specialty |
| **e-Table 17** | Outcomes Associated with Normal Spirometry in PFTs with Restriction |
| **e-Table 18** | Differences in Respiratory Diseases Associated with Restriction in PFTs with Normal Spirometry when Diseases Are Assigned Using the MAP Algorithm versus the Presence of at least One ICD-10 Code |
| **e-Table 19** | Differences in Outcomes Associated with Restriction in PFTs with Normal Spirometry when Normal Spirometry Is Defined with Race-Specific versus Race-Neutral Reference |
| **e-Table 20** | Differences in Outcomes Associated with Restriction in PFTs with Normal Spirometry when Normal Spirometry Is Defined with Race-Specific versus Race-Neutral Reference Equations, by Race |


**Figures**


| | |
|---|---|
| **e-Figure 1** | Flow Diagram for the Inclusion of PFTs in this Study |
| **e-Figure 2** | Persistence of PFT Interpretations with Race-Specific Reference Equations Reference Equations |

### e-Table 1 — STROBE Checklist

| | Item Number | Recommendation | Location |
|---|---|---|---|
| **Title and Abstract** | 1 | (a) Indicate the study's design with a commonly used term in the title or the abstract | Title |
| | | (b) Provide in the abstract an informative and balanced summary of what was done and what was found | Abstract |
| **Introduction** | | | |
| Background/rationale | 2 | Explain the scientific background and rationale for the investigation being reported | Page 4 |
| Objectives | 3 | State specific objectives, including any prespecified hypotheses | Page 4 |
| **Methods** | | | |
| Study design | 4 | Present key elements of study design early in the paper | Page 5 |
| Setting | 5 | Describe the setting, locations, and relevant dates, including periods of recruitment, exposure, follow-up, and data collection | Page 5 |
| Participants | 6 | (a) *Cohort Study*—Give the eligibility criteria, and the sources and methods of selection of participants. Describe methods of follow-up | Page 5 |
| | | (b) *Cohort Study*—For matched studies, give matching criteria and number of exposed and unexposed | NA |
| Variables | 7 | Clearly define all outcomes, exposures, predictors, potential confounders, and effect modifiers. Give diagnostic criteria, if applicable | Pages 5–8 |
| Data sources/measurement | 8 | For each variable of interest, give sources of data and details of methods of assessment (measurement). Describe comparability of assessment methods if there is more than one group | Pages 5–8 |
| Bias | 9 | Describe any efforts to address potential sources of bias | Page 8 |
| Study size | 10 | Explain how the study size was arrived at | Page 5 |
| Quantitative variables | 11 | Explain how quantitative variables were handled in the analyses. If applicable, describe which groupings were chosen and why | NA |
| Statistical Methods | 12 | (a) Describe all statistical methods, including those used to control for confounding | Pages 7–8 |
| | | (b) Describe any methods used to examine subgroups and interactions | Page 7 |
| | | (c) Explain how missing data were addressed | Page 14 |
| | | (d) *Cohort study*—If applicable, explain how loss to follow-up was addressed | Page 5 |
| | | (e) Describe any sensitivity analyses | Page 6 |

**e-Table 1 (Continued) — STROBE Checklist**

| | Item Number | Recommendation | Location |
|---|---|---|---|
| **Results** | | | |
| Participants | 13 | (a) Report numbers of individuals at each stage of study—e.g. numbers potentially eligible, examined for eligibility, confirmed eligible, included in the study, completing follow-up, and analysed | e-Figure 1 |
| | | (b) Give reasons for non-participation at each stage | e-Figure 1 |
| | | (c) Consider use of a flow diagram | e-Figure 1 |
| Descriptive Data | 14 | (a) Give characteristics of study participants (e.g. demographic, clinical, social) and information on exposures and potential confounders | Table 1 |
| | | (b) Indicate number of participants with missing data for each variable of interest | e-Table 11 |
| | | (c) *Cohort study*—Summarize follow-up time (e.g., average and total amount) | Page 8 |
| Outcome Data | 15 | *Cohort study*—Report numbers of outcome events or summary measures over time | Figure 4 |
| Main Results | 16 | (a) Give unadjusted estimates and, if applicable, confounder-adjusted estimates and their precision (e.g., 95% confidence interval). Make clear which confounders were adjusted for and why they were included | Figures 3–4 |
| | | (b) Report category boundaries when continuous variables were categorized | NA |
| | | (c) If relevant, consider translating estimates of relative risk into absolute risk for a meaningful time period | NA |
| Other Analyses | 17 | Report other analyses done—e.g. analyses of subgroups and interactions, and sensitivity analyses | e-Tables 13–20 |
| **Discussion** | | | |
| Key Results | 18 | Summarize key results with reference to study objectives | Pages 10–11 |
| Limitations | 19 | Discuss limitations of the study, taking into account sources of potential bias or imprecision. Discuss both direction and magnitude of any potential bias | Pages 13–14 |
| Interpretation | 20 | Give a cautious overall interpretation of results considering objectives, limitations, multiplicity of analyses, results from similar studies, and other relevant evidence | Page 14 |
| Generalisability | 21 | Discuss the generalisability (external validity) of the study results | Pages 13–14 |
| **Other Information** | | | |
| Funding | 22 | Give the source of funding and the role of the funders for the present study and, if applicable, for the original study on which the present article is based | Title Page |

**e-Table 2 — ICD-10 Codes for Respiratory Diseases**

| Respiratory Disease | ICD-10 Code |
|---|---|
| Asthma | J45.21, J45.22, J45.3, J45.30, J45.31, J45.32, J45.4, J45.40, J45.41, J45.5, J45.50, J45.51, J45.52, J45.9, J45.90, J45.901, J45.902, J45.909, J45.99, J45.990, J45.991, J45.998 |
| Bronchiectasis | E84.0, J47, J47.0, J47.1, J47.9 |
| COPD | J41, J41.0, J41.1, J41.8, J42, J43, J43.0, J43.1, J43.2, J43.8, J43.9, J44, J44.0, J44.1, J44.8, J44.89, J44.9 |
| Chest Wall Disorder | E66.2, I27.1, M40, M40.0, M40.00, M40.03, M40.04, M40.05, M40.1, M40.10, M40.12, M40.13, M40.14, M40.15, M40.2, M40.20, M40.202, M40.203, M40.204, M40.205, M40.209, M40.29, M40.292, M40.293, M40.294, M40.295, M40.299, M40.3, M40.30, M40.35, M40.36, M40.37, M40.4, M40.40, M40.45, M40.46, M40.47, M40.5, M40.50, M40.55, M40.56, M40.57, M41, M41.0, M41.00, M41.02, M41.03, M41.04, M41.05, M41.06, M41.07, M41.08, M41.1, M41.11, M41.112, M41.113, M41.114, M41.115, M41.116, M41.117, M41.119, M41.12, M41.122, M41.123, M41.124, M41.125, M41.126, M41.127, M41.129, M41.2, M41.20, M41.22, M41.23, M41.24, M41.25, M41.26, M41.27, M41.3, M41.30, M41.34, M41.35, M41.4, M41.40, M41.41, M41.42, M41.43, M41.44, M41.45, M41.46, M41.47, M41.5, M41.50, M41.52, M41.53, M41.54, M41.55, M41.56, M41.57, M41.8, M41.80, M41.82, M41.83, M41.84, M41.85, M41.86, M41.87, M41.9, M45, M45.0, M45.1, M45.2, M45.3, M45.4, M45.5, M45.6, M45.7, M45.8, M45.9, M45.A, M45.A0, M45.A1, M45.A2, M45.A3, M45.A4, M45.A5, M45.A6, M45.A7, M45.A8, M45.AB, Q67.5, Q67.6, Q67.7, Q67.8 |
| ILD | D86.0, J60, J61, J62, J62.0, J62.8, J63, J63.0, J63.1, J63.2, J63.3, J63.4, J63.5, J63.6, J64, J65, J67, 67.0, J67.1, J67.2, J67.3, J67.4, J67.5, J67.6, J67.7, J67.8, J67.9, J69, J69.0, J69.1, J69.8, J70.0, J70.1, J70.2, J70.3, J70.4, J82, J82.8, J82.81, J82.82, J82.83, J82.89, J84, J84.0, J84.01, J84.02, J84.03, J84.09, J84.1, J84.10, J84.11, J84.111, J84.112, J84.113, J84.114, J84.115, J84.116, J84.117, J84.17, 84.170, J84.178, J84.2, J84.8, J84.81, J84.82, J84.89, J84.9, M05.1, M05.10, M05.11, M05.111, M05.112, M05.119, M05.12, M05.121, M05.122, M05.129, M05.13, M05.131, M05.132, M05.139, M05.14, M05.141, M05.142, M05.149, M05.15, M05.151, M05.152, M05.159, M05.16, M05.161, M05.162, M05.169, M05.17, M05.171, M05.172, M05.179, M05.19, M32.13, M33.01, M33.11, M33.21, M33.91, M34.81, M35.02 |
| Neuromuscular Disease | G12, G12.0, G12.1, G12.2, G12.20, G12.21, G12.22, G12.23, G12.24, G12.25, G12.29, G12.8, G12.9, G13, G13.0, G13.1, G13.2, G13.8, G35, G61, G61.0, G61.1, G61.8, G61.81, G61.82, G61.89, G61.9, G65, G65.0, G65.1, G65.2, G70, G70.0, G70.00, G70.01, G70.1, G70.2, G70.8, G70.80, G70.81, G70.89, G70.9, G71, G71.0, G71.00, G71.01, G71.02, G71.03, G71.031, G71.032, G71.033, G71.034, G71.0340, G71.0341, G71.0342, G71.0349, G71.035, G71.038, G71.039, G71.09, G71.1, G71.11, G71.12, G71.13, G71.14, G71.19, G71.2, G71.20, G71.21, G71.22, G71.220, G71.228, G71.29, G71.3, G71.8, G71.9, G72, G72.0, G72.1, G72.2, G72.3, G72.4, G72.41, G72.49, G72.8, G72.81, G72.89, G72.9, G73, G73.1, G73.3, G73.7 |

### e-Table 3 — Expressions in Clinic Notes Associated with Respiratory Diseases

| Respiratory Disease | ICD-10 Code |
|---|---|
| Asthma | “asthma”, “hyperresponsive”, “reactive airway disease” “reactive airways disease” |
| Bronchiectasis | “bronchiectasis”, “cystic fibrosis”, “kartagener”, “primary ciliary dyskinesia” |
| Chest Wall Disorder | “ankylosing spondylitis”, “flatback”, “kyphosis”, “kyphoscoliosis”, “lordosis”, “morbid obesity”, “obesity hypoventilation syndrome”, “scoliosis”, |
| COPD | “chronic bronchitis”, “chronic obstructive pulmonary disease”, “copd”, “emphysema” |
| ILD | “desquamative interstitial pneumonia”, “diffuse parenchymal lung disease”, “dpld”, “fibrosing alveolitis”, “hypersensitivity pneumonitis”, “idiopathic interstitial pneumonia”, “idiopathic pulmonary fibrosis”, “interstitial fibrosis”, “interstitial lung disease”, “lymphangioleiomyomatosis”, “lymphoid interstitial pneumonia”, “nonspecific interstitial pneumonia”, “nsip”, “pulmonary fibrosis”, “pulmonary langerhans cell histiocytosis”, “radiation pneumonitis”, “rbild”, “respiratory bronchiolitis interstitial lung disease”, “rheumatoid lung disease”, “sarcoidosis”, “sjogren”, “systemic sclerosis”, “uip”, “usual interstitial pneumonia” |
| Neuromuscular Disease | “amyotrophic lateral sclerosis”, “inclusion body myositis”, “inflammatory polyneuropathy”, “guillain-barre”, “lambert-eaton”, “motor neuron disease”, “muscular atrophy”, “muscular dystrophy”, “multiple sclerosis”, “myasthenia gravis”, “myopathy”, “myotonia”, “myotonic”, “progressive bulbar palsy” |

### e-Table 4 — Expressions in Radiographic Reports Associated with CT Findings

| Finding | Expression |
|---|---|
| Bronchial Wall Thickening | `"(?i)(bronchial|peribronchial|airway)\\s*(wall)?\\s*(thickening|thickened|prominence|prominent|fullness|cuffing|cuffed)"` |
| Emphysema | `"(?i)emphysema|emphysematous|centrilobular|panlobular|paraseptal"` |
| Honeycombing | `"(?i)honeycomb(ing|ed)?|subpleural\\s*cystic\\s*changes"` |
| Reticulation | `"(?i)reticulation|reticular|intralobular\\s*lines|net-like"` |
| Traction Bronchiectasis | `"(?i)traction\\s*bronchiectasis|traction\\s*bronchiolectasis"` |

**e-Table 5 — Expressions in PFT Reports Associated with Adequate and Inadequate Effort**

| Presence of Concern Regarding Patient Effort | Absence of Concern Regarding Patient Effort |
|---|---|
| "efforts appeared inconsistent" | "adequate patient effort" |
| "effort could be stronger" | "best effort throughout" |
| "effort is variable" | "effort appeared good" |
| "efforts variable" | "effort was good" |
| "efforts were marred" | "good effort" |
| "efforts were variable" | "good patient coordination and effort" |
| "effort was variable" | "good patient comprehension and effort" |
| "fair effort" | "good patient effort" |
| "inconsistent effort" | "good pt effort" |
| "poor effort" | "great effort" |
| "questionable effort" | "maximal effort" |
| "questionable patient effort" | "repeatable effort" |
| "suboptimal effort" | |
| "uneven effort" | |
| "variable effort" | |
| "weak effort" | |

### e-Table 6 — Respiratory Reasons for ED Visits

| **Reason** |
|---|
| “airway obstruction” |
| “aspiration” |
| “asthma” |
| “breathing problem” |
| “copd” |
| “cough” |
| “cystic fibrosis” |
| “infleunza” |
| “pneumonia” |
| “respiratory arrest” |
| “respiratory distress” |
| “shortness of breath” |
| “tracheostomy tube change” |
| “uri” |
| “wheezing” |

### e-Table 7 — Characteristics by Patient Sex

| | Male ($n = 33,690$) | Female ($n = 50,196$) |
|---|---|---|
| **Age, yrs** | 60.0 (13.6) | 57.9 (14.1) |
| **Height, cm** | 176.0 (7.6) | 162.1 (6.9) |
| **Race** | | |
| White | 22,846 (71.3%) | 28,013 (58.5%) |
| Black | 6,854 (21.4%) | 16,641 (34.7%) |
| Asian | 1,103 (3.4%) | 1,614 (3.4%) |
| Other | 1,225 (3.8%) | 1,658 (3.5%) |
| **Ethnicity** | | |
| Hispanic | 1,114 (3.4%) | 1,586 (3.2%) |
| Not Hispanic | 31,685 (96.6%) | 47,217 (96.8%) |
| **BMI, kg/m$^2$** | 29.3 (6.3) | 30.4 (8.3) |
| **Pack Years, yr** | 16.9 (37.4) | 9.3 (19.5) |
| **Respiratory Symptoms** | | |
| Cough | 6,714 (48.7%) | 10,421 (51.8%) |
| Dyspnea | 7,165 (52.2%) | 12,359 (61.6%) |
| Wheeze | 5,149 (37.5%) | 7,861 (39.3%) |
| **Respiratory Diseases** | | |
| Asthma | 2,945 (9.0%) | 7,969 (16.5%) |
| Bronchiectasis | 966 (3.0%) | 1,797 (3.7%) |
| Chest Wall Disorder | 215 (0.7%) | 417 (0.9%) |
| Chronic Obstructive Pulmonary Disease | 5,345 (16.4%) | 6,468 (13.4%) |
| Interstitial Lung Disease | 8,926 (27.4%) | 12,927 (26.7%) |
| Neuromuscular Disease | 409 (1.3%) | 618 (1.3%) |
| None | 16,724 (51.3%) | 23,222 (48.0%) |
| **Computed Tomography Findings** | | |
| Bronchial Wall Thickening | 9,619 (35.9%) | 12,086 (31.5%) |
| Emphysema | 11,466 (42.8%) | 14,209 (37.1%) |
| Honeycombing | 4,201 (15.7%) | 3,550 (9.3%) |
| Reticulation | 8,208 (30.7%) | 9,837 (25.7%) |
| Traction Bronchiectasis | 7,047 (26.3%) | 8,600 (22.4%) |
| **Dynamic Lung Volumes, z-score** | | |
| $FEV_1$ | −1.4 (1.5) | −1.2 (1.3) |
| FVC | −1.0 (1.5) | −0.9 (1.3) |
| $FEV_1$/FVC | −0.8 (1.5) | −0.7 (1.4) |
| **Static Lung Volumes, z-score** | | |
| TLC | −1.4 (1.6) | −1.2 (1.7) |
| RV | −0.1 (1.5) | 0.2 (1.3) |
| **Diffusing Capacity, z-score** | | |
| $D_{LCO}$ | −1.9 (2.0) | −1.9 (2.2) |
| **Interpretation** | | |
| Normal | 11,839 (35.1%) | 21,444 (42.7%) |
| Non-Specific | 598 (1.8%) | 1,530 (3.0%) |
| Obstructive | 6,854 (20.3%) | 9,032 (18.0%) |
| Restrictive with Normal Spirometry | 4,721 (14.0%) | 6,640 (13.2%) |
| Restrictive with Abnormal Spirometry | 7,721 (22.9%) | 9,473 (18.9%) |
| Mixed | 1,957 (5.8%) | 2,077 (4.1%) |

Categorical data are presented as No. (%) while continuous data are represented as mean (SD). Missing data have been removed from both calculations. BMI = body mass index; $D_{LCO}$ = diffusing capacity for carbon monoxide; $FEV_1$ = forced expiratory volume in 1 second; FVC = forced vital capacity; RV = residual volume; TLC = total lung capacity.

### e-Table 8 — Characteristics by Patient Race

| | White ($n = 50,859$) | Black ($n = 23,495$) | Asian ($n = 2,717$) | Other ($n = 2,883$) |
|---|---|---|---|---|
| **Age, yrs** | 60.4 (13.7) | 56.6 (13.4) | 54.2 (15.9) | 56.0 (14.9) |
| **Height, cm** | 168.5 (10.1) | 166.7 (9.4) | 162.6 (9.0) | 165.8 (9.9) |
| **Sex** | | | | |
| Male | 22,846 (44.9%) | 6,854 (29.2%) | 1,103 (40.6%) | 1,225 (42.5%) |
| Female | 28,013 (55.1%) | 16,641 (70.8%) | 1,614 (59.4%) | 1,658 (57.5%) |
| **Ethnicity** | | | | |
| Hispanic | 863 (1.7%) | 217 (0.9%) | 19 (0.7%) | 776 (27.9%) |
| Not Hispanic | 49,602 (98.3%) | 23,203 (99.1%) | 2,680 (99.3%) | 2,003 (72.1%) |
| **BMI, kg/m$^2$** | 29.1 (6.8) | 32.5 (8.7) | 25.7 (5.2) | 29.7 (7.0) |
| **Pack Years, yr** | 14.4 (30.6) | 10.3 (27.5) | 3.5 (11.9) | 9.9 (21.2) |
| **Respiratory Symptoms** | | | | |
| Cough | 10,628 (50.3%) | 4,763 (50.7%) | 470 (54.0%) | 609 (53.0%) |
| Dyspnea | 11,553 (55.1%) | 6,097 (64.6%) | 409 (47.5%) | 694 (60.8%) |
| Wheeze | 7,438 (35.4%) | 4,336 (46.4%) | 239 (27.6%) | 489 (42.9%) |
| **Respiratory Diseases** | | | | |
| Asthma | 5,151 (10.3%) | 4,777 (20.5%) | 314 (11.7%) | 405 (14.7%) |
| Bronchiectasis | 2,031 (4.1%) | 415 (1.8%) | 135 (5.0%) | 110 (4.0%) |
| Chest Wall Disorder | 324 (0.7%) | 281 (1.2%) | 7 (0.3%) | 17 (0.6%) |
| Chronic Obstructive Pulmonary Disease | 6,676 (13.4%) | 4,550 (19.5%) | 171 (6.4%) | 251 (9.1%) |
| Interstitial Lung Disease | 14,152 (28.4%) | 5,499 (23.6%) | 832 (31.0%) | 741 (26.8%) |
| Neuromuscular Disease | 690 (1.4%) | 254 (1.1%) | 21 (0.8%) | 20 (0.7%) |
| None | 25,338 (50.8%) | 10,422 (44.7%) | 1,365 (50.9%) | 1,493 (54.1%) |
| **Computed Tomography Findings** | | | | |
| Bronchial Wall Thickening | 13,091 (32.1%) | 7,044 (38.0%) | 659 (32.0%) | 561 (26.8%) |
| Emphysema | 15,463 (37.9%) | 8,503 (45.9%) | 563 (27.3%) | 687 (32.8%) |
| Honeycombing | 5,129 (12.6%) | 1,864 (10.1%) | 311 (15.1%) | 241 (11.5%) |
| Reticulation | 12,041 (29.5%) | 4,474 (24.1%) | 617 (30.0%) | 530 (25.3%) |
| Traction | 10,194 (25.0%) | 4,031 (21.7%) | 534 (25.9%) | 496 (23.7%) |
| **Dynamic Lung Volumes, z-score** | | | | |
| $FEV_1$ | −1.1 (1.4) | −1.9 (1.1) | −1.3 (1.3) | −1.3 (1.4) |
| FVC | −0.7 (1.4) | −1.6 (1.2) | −1.1 (1.4) | −1.0 (1.4) |
| $FEV_1$/FVC | −0.8 (1.4) | −0.8 (1.5) | −0.5 (1.3) | −0.7 (1.4) |
| **Static Lung Volumes, z-score** | | | | |
| TLC | −1.0 (1.6) | −1.8 (1.5) | −1.5 (1.6) | −1.3 (1.6) |
| RV | 0.1 (1.4) | 0.1 (1.3) | −0.0 (1.3) | 0.1 (1.3) |
| **Diffusing Capacity, z-score** | | | | |
| $D_{LCO}$ | −1.7 (2.1) | −2.4 (2.2) | −1.8 (1.9) | −1.7 (2.2) |
| **Interpretation** | | | | |
| Normal | 23,463 (46.1%) | 5,869 (25.0%) | 1,094 (40.3%) | 1,163 (40.3%) |
| Non-Specific | 962 (1.9%) | 919 (3.9%) | 70 (2.6%) | 63 (2.2%) |
| Obstructive | 10,312 (20.3%) | 4,079 (17.4%) | 366 (13.5%) | 505 (17.5%) |
| Restrictive with Normal Spirometry | 6,306 (12.4%) | 3,722 (15.8%) | 393 (14.5%) | 399 (13.8%) |
| Restrictive with Abnormal Spirometry | 8,081 (15.9%) | 7,007 (29.8%) | 686 (25.2%) | 619 (21.5%) |
| Mixed | 1,735 (3.4%) | 1,899 (8.1%) | 108 (4.0%) | 134 (4.6%) |

Categorical data are presented as No. (%) while continuous data are represented as mean (SD). Missing data have been removed from both calculations. BMI = body mass index; $D_{LCO}$ = diffusing capacity for carbon monoxide; $FEV_1$ = forced expiratory volume in 1 second; FVC = forced vital capacity; RV = residual volume; TLC = total lung capacity.

### e-Table 9 — Characteristics by Pulmonary Diagnostic Lab

| | Lab 1 ($n = 55,169$) | Lab 2 ($n = 14,399$) | Lab 3 ($n = 7,835$) | Lab 4 ($n = 3,421$) |
|---|---|---|---|---|
| **Age, yrs** | 58.4 (13.9) | 58.8 (14.0) | 59.5 (14.1) | 61.9 (14.2) |
| **Height, cm** | 167.8 (9.9) | 167.3 (10.0) | 166.6 (9.8) | 169.0 (10.0) |
| **Sex** | | | | |
| Male | 22,717 (41.2%) | 5,383 (37.4%) | 2,890 (36.9%) | 1,436 (42.0%) |
| Female | 32,452 (58.8%) | 9,016 (62.6%) | 4,945 (63.1%) | 1,985 (58.0%) |
| **Race** | | | | |
| White | 34,934 (66.3%) | 6,571 (48.1%) | 4,458 (60.3%) | 2,842 (86.3%) |
| Black | 13,952 (26.5%) | 6,120 (44.8%) | 2,504 (33.9%) | 231 (7.0%) |
| Asian | 1,862 (3.5%) | 436 (3.2%) | 195 (2.6%) | 114 (3.5%) |
| Other | 1,931 (3.7%) | 535 (3.9%) | 233 (3.2%) | 105 (3.2%) |
| **Ethnicity** | | | | |
| Hispanic | 1,676 (3.1%) | 492 (3.5%) | 349 (4.6%) | 103 (3.1%) |
| Not Hispanic | 52,140 (96.9%) | 13,402 (96.5%) | 7,207 (95.4%) | 3,248 (96.9%) |
| **BMI, kg/m$^2$** | 29.8 (7.4) | 30.7 (7.9) | 30.7 (8.1) | 28.8 (6.7) |
| **Pack Years, yr** | 15.0 (31.8) | 9.7 (24.9) | 7.1 (19.1) | — |
| **Respiratory Symptoms** | | | | |
| Cough | 13,842 (49.6%) | 1,584 (55.2%) | 1,709 (54.4%) | — |
| Dyspnea | 15,858 (57.4%) | 1,748 (56.0%) | 1,918 (62.7%) | — |
| Wheeze | 10,323 (37.0%) | 1,278 (46.7%) | 1,409 (45.2%) | — |
| **Respiratory Diseases** | | | | |
| Asthma | 6,325 (11.7%) | 2,804 (21.1%) | 1,283 (17.1%) | 133 (4.1%) |
| Bronchiectasis | 2,072 (3.8%) | 288 (2.2%) | 257 (3.4%) | 24 (0.7%) |
| Chest Wall Disorder | 408 (0.8%) | 121 (0.9%) | 71 (0.9%) | 12 (0.4%) |
| Chronic Obstructive Pulmonary Disease | 6,846 (12.7%) | 2,792 (21.0%) | 1,645 (22.0%) | 122 (3.8%) |
| Interstitial Lung Disease | 18,146 (33.6%) | 2,084 (15.7%) | 518 (6.9%) | 167 (5.2%) |
| Neuromuscular Disease | 739 (1.4%) | 157 (1.2%) | 66 (0.9%) | 26 (0.8%) |
| None | 25,063 (46.4%) | 6,356 (47.9%) | 4,326 (57.7%) | 2,767 (86.2%) |
| **Computed Tomography Findings** | | | | |
| Bronchial Wall Thickening | 14,404 (32.0%) | 3,592 (36.7%) | 2,228 (41.2%) | 615 (26.0%) |
| Emphysema | 17,046 (37.9%) | 4,201 (42.9%) | 2,604 (48.1%) | 833 (35.2%) |
| Honeycombing | 6,340 (14.1%) | 725 (7.4%) | 248 (4.6%) | 129 (5.5%) |
| Reticulation | 13,793 (30.7%) | 2,026 (20.7%) | 1,033 (19.1%) | 446 (18.9%) |
| Traction Bronchiectasis | 12,341 (27.5%) | 1,659 (17.0%) | 711 (13.1%) | 278 (11.8%) |
| **Dynamic Lung Volumes, z-score** | | | | |
| $FEV_1$ | −1.4 (1.4) | −1.5 (1.4) | −1.2 (1.3) | −0.6 (1.3) |
| FVC | −1.0 (1.4) | −1.1 (1.3) | −0.9 (1.3) | −0.2 (1.2) |
| $FEV_1$/FVC | −0.7 (1.4) | −0.9 (1.5) | −0.6 (1.5) | −0.8 (1.2) |
| **Static Lung Volumes, z-score** | | | | |
| TLC | −1.4 (1.7) | −1.3 (1.5) | −0.4 (1.5) | −0.8 (1.4) |
| RV | −0.1 (1.3) | 0.1 (1.3) | 1.1 (1.3) | 0.1 (1.2) |
| **Diffusing Capacity, z-score** | | | | |
| $D_{LCO}$ | −1.9 (2.1) | −2.3 (2.1) | −1.4 (2.4) | −1.3 (1.7) |
| **Interpretation** | | | | |
| Normal | 20,865 (37.8%) | 5,083 (35.3%) | 4,194 (53.5%) | 1,962 (57.4%) |
| Non-Specific | 1,160 (2.1%) | 326 (2.3%) | 561 (7.2%) | 31 (0.9%) |
| Obstructive | 9,724 (17.6%) | 3,292 (22.9%) | 1,618 (20.7%) | 627 (18.3%) |
| Restrictive with Normal Spirometry | 7,933 (14.4%) | 2,048 (14.2%) | 361 (4.6%) | 457 (13.4%) |
| Restrictive with Abnormal Spirometry | 12,535 (22.7%) | 2,886 (20.0%) | 1,007 (12.9%) | 273 (8.0%) |
| Mixed | 2,952 (5.4%) | 764 (5.3%) | 94 (1.2%) | 71 (2.1%) |

Categorical data are presented as No. (%) while continuous data are represented as mean (SD). Missing data have been removed from both calculations. BMI = body mass index; $D_{LCO}$ = diffusing capacity for carbon monoxide; $FEV_1$ = forced expiratory volume in 1 second; FVC = forced vital capacity; RV = residual volume; TLC = total lung capacity.

### e-Table 10 — Characteristics by Referring Specialty

| | **Pulmonology** ($n = 26,753$) | **Primary Care** ($n = 3,580$) |
|---|---|---|
| **Age, yrs** | 60.8 (12.7) | 57.4 (14.9) |
| **Height, cm** | 167.6 (9.9) | 167.6 (9.7) |
| **Sex** | | |
| Male | 10,792 (40.3%) | 1,274 (35.6%) |
| Female | 15,961 (59.7%) | 2,306 (64.4%) |
| **Race** | | |
| White | 17,874 (66.8%) | 1,927 (53.8%) |
| Black | 6,999 (26.2%) | 1,367 (38.2%) |
| Asian | 906 (3.4%) | 165 (4.6%) |
| Other | 974 (3.6%) | 121 (3.4%) |
| **Ethnicity** | | |
| Hispanic | 537 (2.0%) | 86 (2.4%) |
| Not Hispanic | 26,011 (98.0%) | 3,471 (97.6%) |
| **BMI, kg/m$^2$** | 29.8 (7.2) | 29.9 (7.5) |
| **Pack Years, yr** | 13.7 (28.5) | 8.4 (18.2) |
| **Respiratory Symptoms** | | |
| Cough | 6,619 (55.0%) | 718 (55.4%) |
| Dyspnea | 7,169 (59.9%) | 764 (59.5%) |
| Wheeze | 4,759 (39.7%) | 589 (45.8%) |
| **Respiratory Diseases** | | |
| Asthma | 3,519 (13.2%) | 520 (14.5%) |
| Bronchiectasis | 1,294 (4.8%) | 32 (0.9%) |
| Chest Wall Disorder | 194 (0.7%) | 36 (1.0%) |
| Chronic Obstructive Pulmonary Disease | 4,455 (16.7%) | 397 (11.1%) |
| Interstitial Lung Disease | 10,830 (40.6%) | 105 (2.9%) |
| Neuromuscular Disease | 344 (1.3%) | 32 (0.9%) |
| None | 9,509 (35.6%) | 2,578 (72.1%) |
| **Computed Tomography Findings** | | |
| Bronchial Wall Thickening | 7,654 (32.7%) | 781 (36.6%) |
| Emphysema | 9,254 (39.5%) | 850 (39.8%) |
| Honeycombing | 3,732 (15.9%) | 52 (2.4%) |
| Reticulation | 7,970 (34.0%) | 286 (13.4%) |
| Traction Bronchiectasis | 7,301 (31.2%) | 152 (7.1%) |
| **Dynamic Lung Volumes, z-score** | | |
| $FEV_1$ | −1.4 (1.3) | −0.8 (1.3) |
| FVC | −1.1 (1.4) | −0.5 (1.3) |
| $FEV_1$/FVC | −0.7 (1.5) | −0.7 (1.2) |
| **Static Lung Volumes, z-score** | | |
| TLC | −1.4 (1.7) | −0.7 (1.3) |
| RV | −0.1 (1.4) | 0.4 (1.1) |
| **Diffusing Capacity, z-score** | | |
| $D_{LCO}$ | −2.1 (2.1) | −0.9 (1.7) |
| **Interpretation** | | |
| Normal | 9,130 (34.1%) | 2,067 (57.7%) |
| Non-Specific | 625 (2.3%) | 92 (2.6%) |
| Obstructive | 5,134 (19.2%) | 630 (17.6%) |
| Restrictive with Normal Spirometry | 4,114 (15.4%) | 347 (9.7%) |
| Restrictive with Abnormal Spirometry | 6,371 (23.8%) | 345 (9.6%) |
| Mixed | 1,379 (5.2%) | 99 (2.8%) |

Categorical data are presented as No. (%) while continuous data are represented as mean (SD). Missing data have been removed from both calculations. BMI = body mass index; $D_{LCO}$ = diffusing capacity for carbon monoxide; $FEV_1$ = forced expiratory volume in 1 second; FVC = forced vital capacity; RV = residual volume; TLC = total lung capacity.

### e-Table 11 — Missing Data

| | Number of Tests with Observation Missing | Percentage of Tests with Observation Missing |
|---|---|---|
| Race and Ethnicity | 3,932 | 4.7% |
| Pack Years | 47,908 | 57.1% |
| Cough | 49,970 | 59.6% |
| Dyspnea | 50,094 | 59.7% |
| Wheeze | 50,152 | 59.8% |
| Respiratory Disease | 2,927 | 3.5% |
| Computed Tomography | 18,816 | 22.4% |
| Bronchodilator Response | 61,554 | 73.4% |
| Spirometry Effort | 44,318 | 52.8% |
| Residual Volume | 437 | 0.5% |
| Diffusing Capacity for Carbon Monoxide | 5,731 | 6.8% |
| Pulmonary Diagnostic Lab | 3,062 | 3.7% |
| Specialty of Referring Provider | 32,652 | 38.9% |

### e-Table 12 — Characteristics Associated with Restriction in PFTs with Normal Spirometry

| Characteristic | Unadjusted Odds Ratio (95% CI) | Adjusted Odds Ratio (95% CI) |
|---|---|---|
| **Age, yr** | 1.02 (1.01–1.02) | 1.01 (1.01–1.01) |
| **Sex** | | |
| Male | 1.00 (Reference) | 1.00 (Reference) |
| Female | 0.78 (0.75–0.81) | 0.56 (0.52–0.61) |
| **Race** | | |
| White | 1.00 (Reference) | 1.00 (Reference) |
| Black | 2.41 (2.29–2.53) | 1.34 (1.26–1.43) |
| Asian | 1.34 (1.19–1.51) | 1.24 (1.07–1.44) |
| Other | 1.28 (1.14–1.44) | 1.22 (1.05–1.41) |
| **Ethnicity** | | |
| Hispanic | 1.00 (Reference) | 1.00 (Reference) |
| Not Hispanic | 0.97 (0.87–1.09) | 0.83 (0.72–0.96) |
| **BMI, kg/m$^2$** | 1.03 (1.03–1.03) | 1.00 (1.00–1.01) |
| **Pack Years, yr** | 1.00 (1.00–1.00) | 1.00 (1.00–1.00) |
| **Spirometry** | | |
| Bronchodilator Response | 0.45 (0.33–0.59) | 0.39 (0.28–0.52) |
| Adequate Effort | 0.61 (0.37–1.06) | 0.83 (0.45–1.55) |
| **Static Lung Volumes** | | |
| RV, z-score | 0.11 (0.10–0.11) | 0.01 (0.01–0.01) |
| **Diffusing Capacity** | | |
| $D_{LCO}$, z-score | 0.56 (0.55–0.57) | 0.61 (0.60–0.62) |

Adjusted odds ratios are calculated for respiratory symptoms, respiratory disease diagnoses, and chest computed tomography findings, while adjusted hazard ratios are calculated for events. Adjusted odds ratios compare the odds of an outcome among PFTs with normal spirometry, with versus without restriction. Adjusted hazard ratios compare the rate of an event among PFTs with normal spirometry, with versus without restriction. Models are adjusted for patient age, sex, height, $FEV_1$ z-score, FVC z-score, and $FEV_1$/FVC z-score. Abbreviations: CT = computed tomography; ED = emergency department; $FEV_1$ = forced expiratory volume in 1 second; FVC = forced vital capacity.

### e-Table 13 — Outcomes Associated with Restriction in PFTs with Normal Spirometry, by Patient Sex

| | Patient Sex | |
|---|---|---|
| | Male | Female |
| **Outcome** | $n = 16,448$ | $n = 27,798$ |
| **Respiratory Symptoms** | | |
| Cough | 1.40 (1.22–1.61) | 1.15 (1.03–1.29) |
| Dyspnea | 1.28 (1.11–1.48) | 1.13 (1.01–1.27) |
| Wheeze | 0.91 (0.79–1.06) | 0.85 (0.75–0.96) |
| **Respiratory Disease Diagnoses** | | |
| Asthma | 0.74 (0.62–0.87) | 0.82 (0.75–0.90) |
| Broncheictasis | 0.77 (0.57–1.02) | 0.53 (0.43–0.64) |
| Chest Wall Disorder | 0.49 (0.25–0.91) | 0.84 (0.54–1.29) |
| Chronic Obstructive Pulmonary Disease | 1.02 (0.87–1.19) | 0.72 (0.63–0.82) |
| Interstitial Lung Disease | 4.07 (3.69–4.49) | 3.03 (2.81–3.26) |
| Neuromuscular Disorder | 0.71 (0.48–1.04) | 0.65 (0.48–0.88) |
| **Chest Computed Tomography Findings** | | |
| Bronchial Wall Thickening | 0.70 (0.63–0.77) | 0.66 (0.60–0.72) |
| Emphysema | 0.93 (0.84–1.02) | 0.72 (0.67–0.78) |
| Honeycombing | 4.73 (4.08–5.51) | 4.18 (3.63–4.82) |
| Reticulation | 2.93 (2.64–3.26) | 2.43 (2.23–2.64) |
| Traction Bronchiectasis | 3.69 (3.29–4.15) | 3.32 (3.02–3.64) |
| **Event** | | |
| ED Visit with Respiratory Complaint | 1.29 (1.16–1.44) | 1.11 (1.02–1.21) |
| Death from Any Cause | 1.28 (1.14–1.43) | 1.67 (1.49–1.88) |

Adjusted odds ratios are calculated for respiratory symptoms, respiratory disease diagnoses, and chest computed tomography findings, while adjusted hazard ratios are calculated for events. Adjusted odds ratios compare the odds of an outcome among PFTs with normal spirometry, with versus without restriction. Adjusted hazard ratios compare the rate of an event among PFTs with normal spirometry, with versus without restriction. Models are adjusted for patient age, sex, height, $FEV_1$ z-score, FVC z-score, and $FEV_1$/FVC z-score. Abbreviations: CT = computed tomography; ED = emergency department; $FEV_1$ = forced expiratory volume in 1 second; FVC = forced vital capacity.

#### e-Table 14 — Outcomes Associated with Restriction in PFTs with Normal Spirometry, by Patient Race

| | Adjusted Association (95% CI) | | | |
|---|---|---|---|---|
| **Outcome** | **White** ($n = 29,577$) | **Black** ($n = 9,432$) | **Asian** ($n = 1,472$) | **Other** ($n = 1,549$) |
| **Respiratory Symptoms** | | | | |
| Cough | 1.45 (1.30–1.62) | 0.95 (0.81–1.12) | 1.72 (0.98–3.08) | 1.31 (0.80–2.15) |
| Dyspnea | 1.33 (1.19–1.49) | 1.07 (0.91–1.27) | 1.13 (0.63–2.01) | 0.82 (0.50–1.35) |
| Wheeze | 0.85 (0.75–0.96) | 0.91 (0.77–1.08) | 1.48 (0.76–2.85) | 0.82 (0.48–1.40) |
| **Respiratory Disease Diagnoses** | | | | |
| Asthma | 0.63 (0.56–0.71) | 0.89 (0.78–1.00) | 1.09 (0.69–1.70) | 0.65 (0.43–0.97) |
| Broncheictasis | 0.59 (0.48–0.71) | 0.74 (0.50–1.09) | 0.40 (0.18–0.86) | 0.63 (0.16–2.05) |
| Chest Wall Disorder | 0.57 (0.36–0.88) | 1.15 (0.55–2.44) | 3.67 (0.24–205.25) | 0.50 (0.02–6.18) |
| Chronic Obstructive Pulmonary Disease | 0.82 (0.72–0.94) | 0.71 (0.60–0.83) | 1.17 (0.55–2.47) | 1.01 (0.53–1.89) |
| Interstitial Lung Disease | 4.00 (3.71–4.31) | 2.86 (2.53–3.24) | 2.97 (2.10–4.24) | 2.84 (2.04–3.97) |
| Neuromuscular Disorder | 0.58 (0.42–0.77) | 1.04 (0.68–1.59) | 0.24 (0.01–1.56) | 1.07 (0.14–5.94) |
| **Chest Computed Tomography Findings** | | | | |
| Bronchial Wall Thickening | 0.64 (0.59–0.70) | 0.72 (0.64–0.81) | 0.60 (0.41–0.87) | 0.87 (0.59–1.28) |
| Emphysema | 0.78 (0.72–0.84) | 0.75 (0.67–0.84) | 0.51 (0.34–0.77) | 0.82 (0.57–1.19) |
| Honeycombing | 4.73 (4.19–5.34) | 4.06 (3.18–5.20) | 4.27 (2.22–8.65) | 2.42 (1.16–5.16) |
| Reticulation | 3.00 (2.76–3.25) | 1.92 (1.67–2.21) | 3.49 (2.31–5.32) | 2.26 (1.54–3.33) |
| Traction Bronchiectasis | 3.62 (3.32–3.95) | 3.05 (2.60–3.58) | 4.65 (2.91–7.56) | 3.69 (2.40–5.74) |
| **Event** | | | | |
| ED Visit with Respiratory Complaint | 1.32 (1.21–1.45) | 0.92 (0.83–1.02) | 1.31 (0.83–2.07) | 1.67 (1.13–2.47) |
| Death from Any Cause | 1.54 (1.40–1.69) | 1.31 (1.11–1.55) | 1.19 (0.60–2.37) | 1.26 (0.67–2.37) |

Adjusted odds ratios are calculated for respiratory symptoms, respiratory disease diagnoses, and chest computed tomography findings, while adjusted hazard ratios are calculated for events. Adjusted odds ratios compare the odds of an outcome among PFTs with normal spirometry, with versus without restriction. Adjusted hazard ratios compare the rate of an event among PFTs with normal spirometry, with versus without restriction. Models are adjusted for patient age, sex, height, $FEV_1$ z-score, FVC z-score, and $FEV_1$/FVC z-score. Abbreviations: CT = computed tomography; ED = emergency department; $FEV_1$ = forced expiratory volume in 1 second; FVC = forced vital capacity.

**e-Table 15 — Outcomes Associated with Restriction in PFTs with Normal Spirometry, by Pulmonary Diagnostic Lab**

| | Adjusted Association (95% CI) | | | |
|---|---|---|---|---|
| **Outcome** | **Lab 1 ($n = 28,598$)** | **Lab 2 ($n = 7,080$)** | **Lab 3 ($n = 4,424$)** | **Lab 4 ($n = 2,411$)** |
| **Respiratory Symptoms** | | | | |
| Cough | 1.33 (1.21–1.47) | 1.04 (0.78–1.39) | 0.90 (0.66–1.23) | — |
| Dyspnea | 1.22 (1.11–1.35) | 1.34 (1.01–1.77) | 1.13 (0.82–1.58) | — |
| Wheeze | 0.88 (0.79–0.98) | 1.05 (0.77–1.42) | 0.95 (0.70–1.31) | — |
| **Respiratory Disease Diagnoses** | | | | |
| Asthma | 0.80 (0.72–0.89) | 0.83 (0.71–0.98) | 1.00 (0.71–1.37) | 0.79 (0.39–1.53) |
| Broncheictasis | 0.52 (0.43–0.63) | 0.64 (0.41–0.99) | 1.29 (0.63–2.45) | 0.96 (0.17–4.72) |
| Chest Wall Disorder | 0.72 (0.47–1.10) | 0.46 (0.17–1.12) | — | — |
| Chronic Obstructive Pulmonary Disease | 0.92 (0.81–1.05) | 0.84 (0.68–1.04) | 0.96 (0.63–1.41) | 1.86 (0.62–5.46) |
| Interstitial Lung Disease | 3.10 (2.89–3.32) | 3.44 (2.88–4.10) | 1.92 (1.31–2.77) | 3.06 (1.75–5.41) |
| Neuromuscular Disorder | 0.54 (0.41–0.72) | 1.30 (0.76–2.22) | 0.76 (0.18–2.30) | 0.64 (0.13–2.55) |
| **Chest Computed Tomography Findings** | | | | |
| Bronchial Wall Thickening | 0.66 (0.61–0.72) | 0.82 (0.69–0.98) | 0.88 (0.65–1.19) | 0.81 (0.54–1.21) |
| Emphysema | 0.83 (0.77–0.89) | 0.80 (0.68–0.94) | 0.67 (0.49–0.89) | 0.90 (0.63–1.30) |
| Honeycombing | 4.13 (3.67–4.66) | 3.87 (2.83–5.35) | 3.98 (2.35–6.68) | 3.11 (1.58–6.31) |
| Reticulation | 2.56 (2.36–2.77) | 2.16 (1.80–2.60) | 1.44 (1.02–2.02) | 2.38 (1.62–3.50) |
| Traction Bronchiectasis | 3.32 (3.05–3.62) | 3.00 (2.44–3.68) | 2.54 (1.75–3.67) | 1.86 (1.16–2.99) |
| **Event** | | | | |
| ED Visit with Respiratory Complaint | 1.29 (1.19–1.40) | 1.21 (1.02–1.43) | 1.30 (1.04–1.62) | 1.48 (0.88–2.48) |
| Death from Any Cause | 1.38 (1.26–1.52) | 1.28 (0.99–1.66) | 1.67 (1.17–2.38) | 1.04 (0.51–2.12) |

Adjusted odds ratios are calculated for respiratory symptoms, respiratory disease diagnoses, and chest computed tomography findings, while adjusted hazard ratios are calculated for events. Adjusted odds ratios compare the odds of an outcome among PFTs with normal spirometry, with versus without restriction. Adjusted hazard ratios compare the rate of an event among PFTs with normal spirometry, with versus without restriction. Models are adjusted for patient age, sex, height, $FEV_1$ z-score, FVC z-score, and $FEV_1$/FVC z-score. Abbreviations: CT = computed tomography; ED = emergency department; $FEV_1$ = forced expiratory volume in 1 second; FVC = forced vital capacity.

### e-Table 16 — Outcomes Associated with Restriction in PFTs with Normal Spirometry, by Referring Specialty

| | Adjusted Association (95% CI) | |
|---|---|---|
| **Outcome** | **Pulmonology ($n = 13,584$)** | **Primary Care ($n = 2,456$)** |
| **Respiratory Symptoms** | | |
| Cough | 1.29 (1.12–1.49) | 1.28 (0.82–2.01) |
| Dyspnea | 1.15 (1.00–1.33) | 1.76 (1.09–2.90) |
| Wheeze | 0.86 (0.74–1.00) | 1.19 (0.76–1.87) |
| **Respiratory Disease Diagnoses** | | |
| Asthma | 0.72 (0.62–0.83) | 1.19 (0.83–1.70) |
| Broncheictasis | 0.46 (0.36–0.58) | 0.59 (0.09–2.38) |
| Chest Wall Disorder | 0.61 (0.31–1.16) | 0.89 (0.13–3.93) |
| Chronic Obstructive Pulmonary Disease | 0.77 (0.65–0.90) | 0.71 (0.40–1.21) |
| Interstitial Lung Disease | 3.24 (2.96–3.56) | 2.81 (1.38–5.72) |
| Neuromuscular Disorder | 0.45 (0.30–0.67) | 0.86 (0.19–3.00) |
| **Chest Computed Tomography Findings** | | |
| Bronchial Wall Thickening | 0.60 (0.54–0.67) | 0.76 (0.52–1.08) |
| Emphysema | 0.85 (0.77–0.94) | 0.86 (0.60–1.22) |
| Honeycombing | 3.84 (3.32–4.45) | 2.02 (0.76–5.21) |
| Reticulation | 2.64 (2.37–2.93) | 0.89 (0.55–1.39) |
| Traction Bronchiectasis | 3.35 (3.00–3.74) | 1.38 (0.75–2.49) |
| **Event** | | |
| ED Visit with Respiratory Complaint | 1.15 (1.02–1.29) | 1.19 (0.86–1.64) |
| Death from Any Cause | 1.68 (1.45–1.95) | 1.42 (0.70–2.87) |

Adjusted odds ratios are calculated for respiratory symptoms, respiratory disease diagnoses, and chest computed tomography findings, while adjusted hazard ratios are calculated for events. Adjusted odds ratios compare the odds of an outcome among PFTs with normal spirometry, with versus without restriction. Adjusted hazard ratios compare the rate of an event among PFTs with normal spirometry, with versus without restriction. Models are adjusted for patient age, sex, height, $FEV_1$ z-score, FVC z-score, and $FEV_1$/FVC z-score. Abbreviations: CT = computed tomography; ED = emergency department; $FEV_1$ = forced expiratory volume in 1 second; FVC = forced vital capacity.

**e-Table 17 — Outcomes Associated with Normal Spirometry in PFTs with Restriction**

| Outcome | Adjusted Association (95% CI) |
|---|---|
| **Respiratory Symptoms** | |
| Cough | 0.90 (0.82–0.98) |
| Dyspnea | 0.61 (0.56–0.67) |
| Wheeze | 0.50 (0.46–0.55) |
| **Respiratory Disease Diagnoses** | |
| Asthma | 0.62 (0.57–0.67) |
| Broncheictasis | 0.67 (0.57–0.79) |
| Chest Wall Disorder | 0.30 (0.21–0.41) |
| Chronic Obstructive Pulmonary Disease | 0.37 (0.34–0.41) |
| Interstitial Lung Disease | 1.80 (1.70–1.90) |
| Neuromuscular Disorder | 0.98 (0.77–1.26) |
| **Chest Computed Tomography Findings** | |
| Bronchial Wall Thickening | 0.57 (0.53–0.61) |
| Emphysema | 0.71 (0.66–0.75) |
| Honeycombing | 1.54 (1.43–1.66) |
| Reticulation | 1.79 (1.68–1.90) |
| Traction Bronchiectasis | 1.56 (1.46–1.66) |
| **Events** | |
| ED Visit with Respiratory Complaint | 1.01 (1.01–1.02) |
| Death from Any Cause | 1.06 (1.06–1.07) |

Adjusted odds ratios are calculated for respiratory symptoms, respiratory disease diagnoses, and chest computed tomography findings, while adjusted hazard ratios are calculated for events. Adjusted odds ratios compare the odds of an outcome among PFTs with restriction, with and without normal spirometry. Adjusted hazard ratios compare the rate of an event among PFTs with restriction, with and without normal spirometry. Logistic regression and Cox proportional hazards models are adjusted for patient age, sex, height, and total lung capacity z-score. Abbreviations: ED = emergency department; PFT = pulmonary function test.

**e-Table 18 — Differences in Respiratory Diseases Associated with Restriction in PFTs with Normal Spirometry when Diseases Are Assigned Using the MAP Algorithm versus the Presence of at least One ICD-10 Code**

| | Adjusted Odds Ratio (95% CI) | |
|---|---|---|
| **Respiratory Disease** | **MAP Algorithm** | **≥ 1 ICD Code** |
| Asthma | 0.79 (0.73–0.86) | 0.86 (0.81–0.91) |
| Broncheictasis | 0.59 (0.50–0.69) | 0.99 (0.90–1.09) |
| Chest Wall Disorder | 0.68 (0.47–0.97) | 0.81 (0.66–0.99) |
| Chronic Obstructive Pulmonary Disease | 0.82 (0.74–0.90) | 0.79 (0.74–0.85) |
| Interstitial Lung Disease | 3.40 (3.20–3.60) | 2.81 (2.66–2.96) |
| Neuromuscular Disorder | 0.68 (0.54–0.86) | 0.94 (0.80–1.09) |

Adjusted odds ratios compare the odds of respiratory disease diagnosis among PFTs with normal spirometry, with and without restriction. Models are adjusted for patient age, sex, height, and total lung capacity z-score. Abbreviations: ICD = International Classification of Disease; MAP = Multimodal Automated Phenotyping.

### e-Table 19 — Differences in Outcomes Associated with Restriction in PFTs with Normal Spirometry with Race-Specific versus Race-Neutral Reference Equations

| | Adjusted Association (95% CI) | |
|---|---|---|
| **Outcome** | **Race-Neutral Equations** | **Race-Specific Equations** |
| **Respiratory Symptoms** | | |
| Cough | 1.25 (1.15–1.36) | 1.22 (1.12–1.32) |
| Dyspnea | 1.20 (1.10–1.31) | 1.44 (1.32–1.57) |
| Wheeze | 0.87 (0.79–0.96) | 1.09 (1.00–1.20) |
| **Respiratory Disease Diagnoses** | | |
| Asthma | 0.79 (0.73–0.86) | 1.00 (0.93–1.08) |
| Broncheictasis | 0.59 (0.50–0.69) | 0.54 (0.46–0.63) |
| Chest Wall Disorder | 0.68 (0.47–0.97) | 0.82 (0.59–1.12) |
| Chronic Obstructive Pulmonary Disease | 0.82 (0.74–0.90) | 1.13 (1.03–1.24) |
| Interstitial Lung Disease | 3.40 (3.20–3.60) | 2.76 (2.61–2.92) |
| Neuromuscular Disorder | 0.68 (0.54–0.86) | 0.74 (0.59–0.92) |
| **Chest Computed Tomography Findings** | | |
| Bronchial Wall Thickening | 0.67 (0.63–0.72) | 0.75 (0.70–0.80) |
| Emphysema | 0.79 (0.74–0.84) | 0.98 (0.92–1.04) |
| Honeycombing | 4.47 (4.03–4.96) | 3.85 (3.49–4.25) |
| Reticulation | 2.63 (2.46–2.81) | 2.24 (2.11–2.39) |
| Traction Bronchiectasis | 3.48 (3.24–3.74) | 3.07 (2.87–3.30) |
| **Event** | | |
| ED Visit with Respiratory Complaint | 1.18 (1.11–1.26) | 1.51 (1.42–1.60) |
| Death from Any Cause | 1.45 (1.34–1.57) | 1.38 (1.28–1.50) |

Normal spirometry is defined using GLI GLobal reference equations for race-neutral reference equations, and defined using GLI 2012 reference equations for race-specific reference equations. Adjusted odds ratios are calculated for respiratory symptoms, respiratory disease diagnoses, and chest computed tomography findings, while adjusted hazard ratios are calculated for events. Adjusted odds ratios compare the odds of an outcome among PFTs with normal spirometry, with versus without restriction. Adjusted hazard ratios compare the rate of an event among PFTs with normal spirometry, with versus without restriction. Logistic regression and Cox proportional hazards models adjust for patient age, sex, height, $FEV_1$ z-score, FVC z-score, and $FEV_1$/FVC z-score. For race-neutral reference equations, adjustments are made using z-scores calculated with GLI Global equations, while for race-specific reference equations, adjustments are made using z-scores calculated with GLI 2012 equations. Abbreviations: CT = computed tomography; ED = emergency department; $FEV_1$ = forced expiratory volume in 1 second; FVC = forced vital capacity; GLI = Global Lung Function Initiative.

### e-Table 20 — Outcomes Associated with Restriction in PFTs with Normal Spirometry with Race-Specific Reference Equations, by Race

| | Adjusted Association (95% CI) | | | |
|---|---|---|---|---|
| | White Patients | | Black Patients | |
| **Outcome** | **Race-Neutral** | **Race-Specific** | **Race-Neutral** | **Race-Specific** |
| **Respiratory Symptoms** | | | | |
| Cough | 1.45 (1.30–1.62) | 1.52 (1.33–1.74) | 0.95 (0.81–1.12) | 0.94 (0.82–1.08) |
| Dyspnea | 1.33 (1.19–1.49) | 1.50 (1.31–1.71) | 1.07 (0.91–1.27) | 0.99 (0.86–1.15) |
| Wheeze | 0.85 (0.75–0.96) | 0.92 (0.79–1.07) | 0.91 (0.77–1.08) | 0.82 (0.71–0.94) |
| **Respiratory Diseases** | | | | |
| Asthma | 0.63 (0.56–0.71) | 0.61 (0.53–0.71) | 0.89 (0.78–1.00) | 0.84 (0.76–0.94) |
| Bronchiectasis | 0.59 (0.48–0.71) | 0.60 (0.48–0.74) | 0.74 (0.50–1.09) | 0.83 (0.58–1.18) |
| Chest Wall Disorder | 0.57 (0.36–0.88) | 0.60 (0.35–0.99) | 1.15 (0.55–2.44) | 1.33 (0.74–2.47) |
| Chronic Obstructive Pulmonary Disease | 0.82 (0.72–0.94) | 0.92 (0.80–1.07) | 0.71 (0.60–0.83) | 0.69 (0.60–0.79) |
| Interstitial Lung Disease | 4.00 (3.71–4.31) | 3.98 (3.67–4.31) | 2.86 (2.53–3.24) | 2.91 (2.60–3.26) |
| Neuromuscular Disorder | 0.58 (0.42–0.77) | 0.59 (0.41–0.84) | 1.04 (0.68–1.59) | 0.96 (0.66–1.41) |
| **Computed Tomography Findings** | | | | |
| Bronchial Wall Thickening | 0.64 (0.59–0.70) | 0.60 (0.54–0.66) | 0.72 (0.64–0.81) | 0.69 (0.62–0.77) |
| Emphysema | 0.78 (0.72–0.84) | 0.82 (0.75–0.89) | 0.75 (0.67–0.84) | 0.73 (0.66–0.81) |
| Honeycombing | 4.73 (4.19–5.34) | 4.31 (3.80–4.90) | 4.06 (3.18–5.20) | 4.32 (3.49–5.39) |
| Reticulation | 3.00 (2.76–3.25) | 2.82 (2.58–3.09) | 1.92 (1.67–2.21) | 2.05 (1.81–2.32) |
| Traction Bronchiectasis | 3.62 (3.32–3.95) | 3.39 (3.08–3.72) | 3.05 (2.60–3.58) | 3.33 (2.89–3.85) |
| **Events** | | | | |
| ED Visit with Respiratory Complaint | 1.32 (1.21–1.45) | 1.41 (1.27–1.56) | 0.92 (0.83–1.02) | 0.94 (0.86–1.02) |
| Death from Any Cause | 1.54 (1.40–1.69) | 1.57 (1.41–1.74) | 1.31 (1.11–1.55) | 1.29 (1.11–1.48) |

Normal spirometry is defined using GLI Global reference equations for race-neutral reference equations, and defined using GLI 2012 reference equations for race-specific reference equations. Adjusted odds ratios are calculated for respiratory symptoms, respiratory disease diagnoses, and chest computed tomography findings, while adjusted hazard ratios are calculated for events. Adjusted odds ratios compare the odds of an outcome among PFTs with normal spirometry, with versus without restriction. Adjusted hazard ratios compare the rate of an event among PFTs with normal spirometry, with versus without restriction. Logistic regression and Cox proportional hazards models adjust for patient age, sex, height, $FEV_1$ z-score, FVC z-score, and $FEV_1$/FVC z-score. For race-neutral reference equations, adjustments are made using z-scores calculated with GLI Global equations, while for race-specific reference equations, adjustments are made using z-scores calculated with GLI 2012 equations. Abbreviations: CT = computed tomography; ED = emergency department; $FEV_1$ = forced expiratory volume in 1 second; FVC = forced vital capacity; GLI = Global Lung Function Initiative.

e-Figure 1 — Flow Diagram for the Inclusion of PFTs in this Study

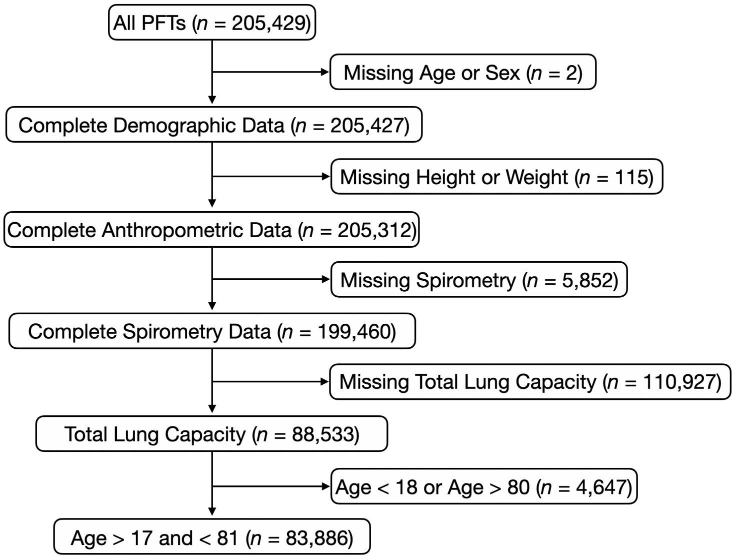

### e-Figure 2 — Race-Specific Reference Equations and the Persistence of PFT Interpretations

A

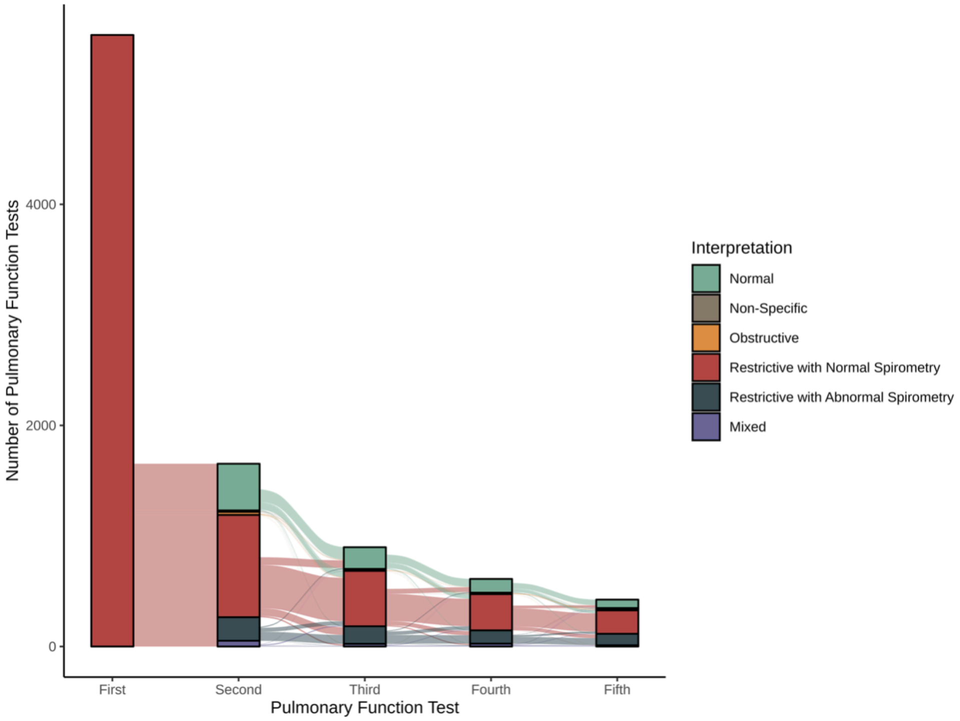


B

**Second Pulmonary Function Test** (columns); **First Pulmonary Function Test** (rows)

| First \ Second | Normal | Non-Specific | Obstructive | Restrictive with Normal Spirometry | Restrictive with Abnormal Spirometry | Mixed |
|---|---|---|---|---|---|---|
| Normal | **76.8%** | 1.8% | 7.6% | 9.8% | 3.0% | 0.9% |
| Non-Specific | 22.3% | **32.7%** | 12.5% | 4.9% | 23.8% | 3.8% |
| Obstructive | 10.1% | 1.3% | **81.4%** | 1.2% | 0.9% | 5.1% |
| Restrictive with Normal Spirometry | 19.4% | 0.6% | 1.2% | **63.1%** | 12.8% | 2.8% |
| Restrictive with Abnormal Spirometry | 2.8% | 2.9% | 0.9% | 8.4% | **80.5%** | 4.6% |
| Mixed | 2.8% | 1.6% | 15.6% | 7.6% | 16.3% | **56.1%** |
